# BinomiRare: A carriers-only test for association of rare genetic variants with a binary outcome for mixed models and any case-control proportion

**DOI:** 10.1101/2021.01.08.21249450

**Authors:** Tamar Sofer, Jiwon Lee, Nuzulul Kurniansyah, Deepti Jain, Cecelia A. Laurie, Stephanie M. Gogarten, Matthew P. Conomos, Ben Heavner, Yao Hu, Charles Kooperberg, Jeffrey Haessler, Ramachandran S. Vasan, L. Adrienne Cupples, Brandon J. Coombes, Amanda Seyerle, Sina A. Gharib, Han Chen, Jeffrey R. O’Connell, Man Zhang, Daniel Gottlieb, Bruce M. Psaty, W.T. Longstreth, Jerome I. Rotter, Kent D. Taylor, Stephen S. Rich, Xiuqing Guo, Eric Boerwinkle, Alanna C. Morrison, James S. Pankow, Andrew D. Johnson, Nathan Pankratz, NHLBI Trans-Omics for Precision Medicine (TOPMed) Consortium, Alex P. Reiner, Susan Redline, Nicholas L. Smith, Kenneth M. Rice, Elizabeth D. Schifano

## Abstract

Whole genome and exome sequencing studies have become increasingly available and are being used to identify rare genetic variants associated with health and disease outcomes. Investigators routinely use mixed models to account for genetic relatedness or other clustering variables (e.g. family or household) when testing genetic associations. However, no existing tests of the association of a rare variant association with a binary outcome in the presence of correlated data controls the Type 1 error where there are (1) few carriers of the rare allele, (2) a small proportion of cases relative to controls, and (3) covariates to adjust for. Here, we address all three issues in developing the carriers-only test framework for testing rare variant association with a binary trait. In this framework, we estimate outcome probabilities under the null hypothesis, and then use them, within the carriers, to test variant associations. We extend the BinomiRare test, which was previously proposed for independent observations, and develop the Conway-Maxwell-Poisson (CMP) test, and study their properties in simulations. We show that the BinomiRare test always controls the type 1 error, while the CMP test sometimes does not. We then use the BinomiRare test to test the association of rare genetic variants in target genes with small vessel disease stroke, short sleep, and venous thromboembolism, in whole-genome sequence data from the Trans-Omics for Precision Medicine program.

## Introduction

Whole-genome and exome-sequencing studies are becoming increasingly available to public health researchers, for example, from NHLBI’s Trans-Omics for Precision Medicine (TOPMed) program (1), NHGRI’s Centers for Common Disease Genetics (CCDG), and the UK Biobank (2). As most variant in sequencing datasets are rare, researchers may be interested in using such datasets for detecting rare-variant associations, genome-wide or in a genomic region of interest. They may also seek to confirm suggested associations from other studies or populations, or to assess pathogenicity in large population-based studies of rare variant alleles reported from small family-based studies. For example, Amininejad et al. (3) studied the association of genetic variants within genes associated with monogenic immunodeficiency disorders with Crohn’s Disease. Wright et al. (4) assessed the pathogenicity and penetrance of rare variants identified in clinical studies, in the population-based UK Biobank. Tuijnenburg et al. (5) studied rare genetic variants within *NGKB1* for association with primary immunodeficiency disease. Do et al. (6) studied risk of myocardial infarction in carriers of rare *LDLR* and *APOA5* alleles. Kendall et al.(7) studied cognitive outcomes in carriers of rare copy number variants. These studies demonstrate that there is an interest in testing single rare genetic variant associations with a wide range of health outcomes, including binary outcomes such as disease or affection status.

Testing rare variant associations with binary traits is challenging. It was previously shown that likelihood-based tests such as the Wald, Score, and likelihood ratio tests poorly control Type 1 error when testing for rare variant associations with a binary trait (8; 9). The Score test performance depends on the case-control ratio, and for rare variants, even a small imbalance causes “inflation” (i.e. too many false positive results). A few approaches have been used previously to study rare variant associations in a set of unrelated individuals. Amininejad et al. used a permutation approach to test for association of rare genetic variants with Crohn’s Disease. Wright et al. used Fisher’s exact test. While it is possible to adjust for covariates in the permutation approach and when using Fisher’s exact test to some extent through stratification (10), they do not have the full flexibility of covariate adjustment of a generalized linear model; i.e. they still require the identification of distinct groups in which no additional adjustment is required. Further, permutation tests may also be computationally intensive if low p-values are desired, because the number of required permutations may be large, although there are ways to reduce this computational burden (10). Alternatively, Tuijnenburg et al. used a method called BeviMed (11), implementing a Bayesian model to estimate posterior disease probabilities. The BinomiRare test has also been proposed as a powerful method to test for rare variant associations that can account for covariates (9). The BinomiRare test uses standard methods to compute the disease probabilities in the entire dataset, under the null hypothesis of no association between a specific genetic variant and the binary outcome. Then, for each specific genetic variant, it uses the estimated probabilities in the variant carriers to test the hypothesis that the disease probabilities under the null are the true outcome probabilities in the carriers. The null hypothesis is rejected if the number of carriers with the outcome is inconsistent with their outcome probabilities. However, the previously published version of this method assumed the sample contains only unrelated individuals. Currently, there is no single-variant test that is generally appropriate for testing rare variants when individuals are correlated, e.g., due to known or cryptic genetic relatedness. Notably, the saddlepoint approximation to compute p-values (henceforth SPA; (12)) was first developed to improve the calibration of the Score test when there is case-control imbalance, and then extended in the SAIGE framework for the settings where related individuals are used (13). However, it does not reliably control the Type I error rate when the number of carriers of the rare variants is very small (i.e. tens of individuals; (14)). Therefore, there is a need for a statistical test that is well-calibrated when the number of carriers is low, individuals are potentially related, and there is case-control imbalance.

The previously published version of BinomiRare test (14) is useful in the presence of case-control imbalance, allows for covariate adjustment, controls the Type I error rate for any number of carriers, and can also be used when combining heterogeneous studies and here, we expand its framework for testing rare variant associations when study individuals are correlated. We developed two tests: first, we extended the BinomiRare test to the mixed models setting by applying it on conditional probabilities computed with a mixed model, rather than on marginal probabilities. Second, we developed the Conway-Maxwell-Poisson (CMP) test, which follows the carriers-only framework by using estimated (conditional) disease probabilities like the BinomiRare. For a given rare variant it uses the estimated disease probabilities in the variant carriers to fit the parameters of the CMP distribution, under the null. It then tests whether the observed number of carriers with the outcome is consistent with this distribution. We study these tests using synthetic simulations with varying outcome probabilities, variant allele frequencies, and strengths of correlation between individuals due to genetic relatedness, and also in realistic simulation studies, using real phenotypes and WGS-based variant call set from the TOPMed program. We finally apply the BinomiRare test to test rare variant associations in known disease causing genes for specific disorders: the *NOTCH3* gene and small vessel disease (SVD)ischemic stroke; the *DEC2* (also known as *BHLHE41*) gene and “short sleep”, and the *F5* gene and venous thromboembolism (VTE).

## Methods

### Statistical approach

Let *D*_*i*_ be an indicator of the disease, or another binary outcome, of participant *i*, with value 1 if the person is affected and 0 otherwise, where *i* = 1, …, *n* and the *n* individuals may be correlated. Let ***x***_*i*_ be a *p* × 1 vector of covariate values for the *i*th participant, and *g*_*i*_ be their count of minor alleles for a specified genetic variant. Under the logistic disease model for correlated data:

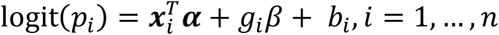

with *p*_*i*_ = pr (*D*_*i*_ = 1|***x***_*i*_, *g*_*i*_, *b*_*i*_) is the conditional outcome probability in the sample (regardless of the population probability), and *b*_*i*_ is the *i* th entry of the vector 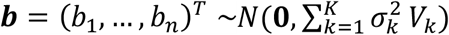 of correlated random effects with possibly *K* variance components 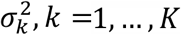 and *V*_*k*_ modelling the correlation structure corresponding to a particular source of correlation. While the methods proposed here can be applied for an arbitrary *K* ≥ 1, we simplify presentation by focusing on the scenario of a single correlation matrix modeling genetic relatedness, possibly cryptic, so that 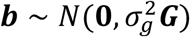 with ***G*** being any genetic relationship matrix (GRM), or possibly kinship matrix, and 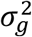 is the corresponding variance component.

We assume that the genetic variant is rare, so that the minor allele frequency (MAF) is low and that carriers of the minor allele are overwhelmingly heterozygotes. While having homozygotes does not invalidate our approach, it also does not increase statistical power. Our carriers-only approach first estimates a disease probability for each individual in the sample under the null hypothesis of no association between the genetic variant and disease status, i.e. under the assumption that *β* = 0 by not including any variant of interest in the regression (step 1, demonstrated in Figure 1), and then considers carriers of the rare variant, testing whether the number of diseased carriers is consistent with their estimated disease probabilities (step 2, demonstrated in Figure 2).

**Figure 1:**
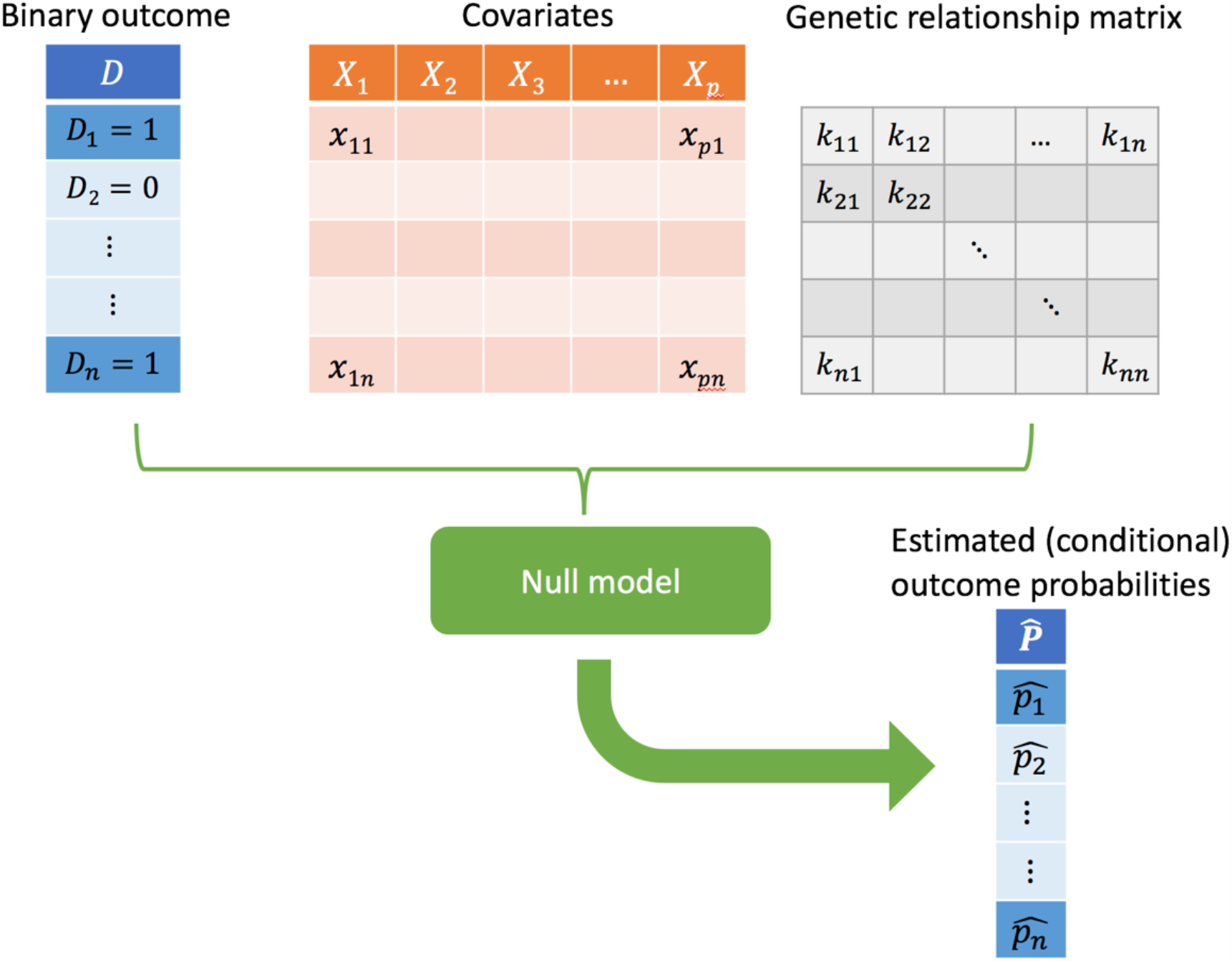
Step 1 of testing genetic association using the carriers-only tests framework. A “null model” of association between the binary outcomes and covariates of interest is fitted, accounting for genetic relationship. Then, estimated conditional outcome probabilities are extracted to be used in the testing step.

**Figure 2:**
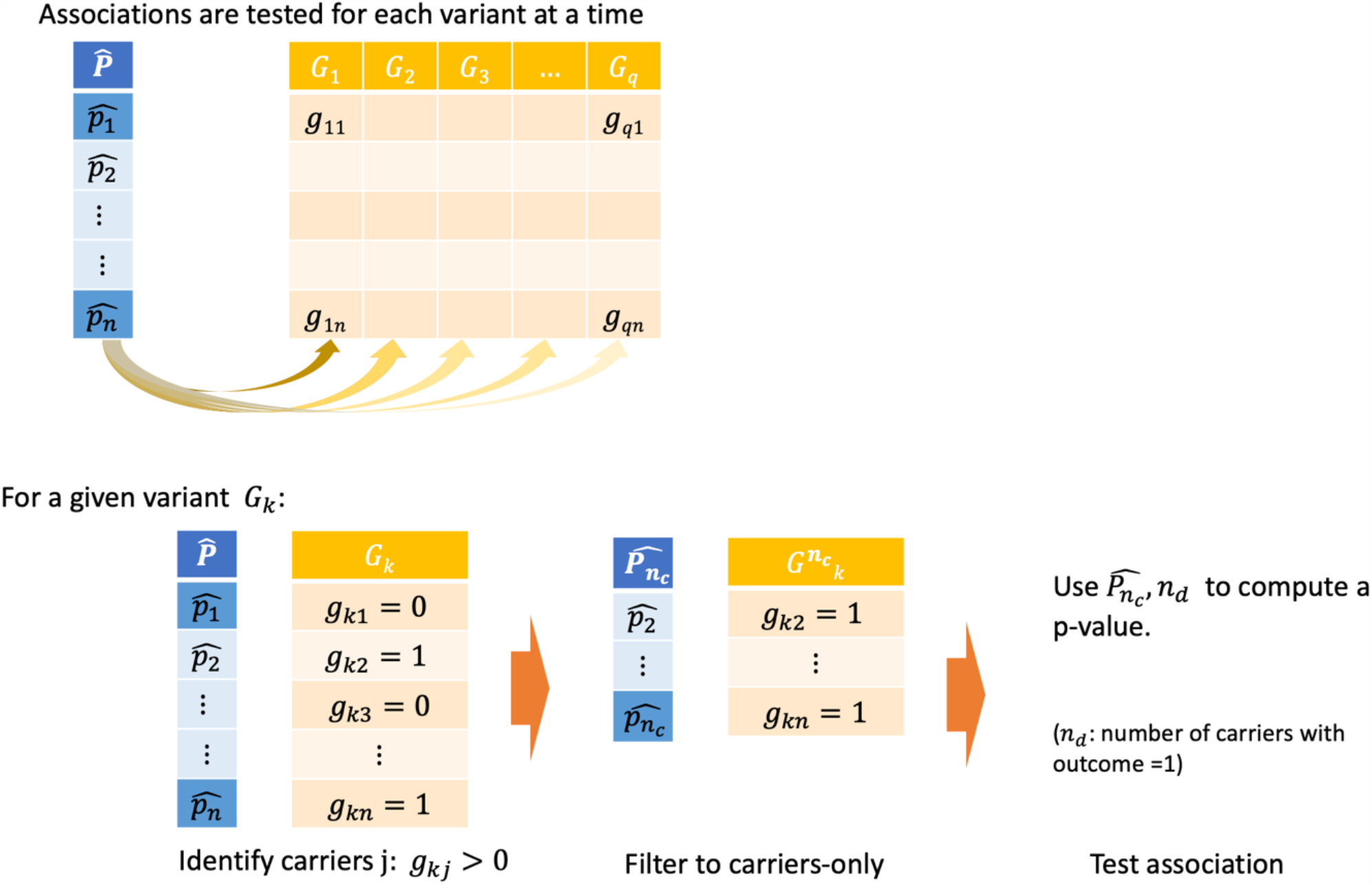
Step 2 of testing genetic associations using the carriers-only tests framework. Based on estimated outcome probabilities, variants are inspected one at a time. For a given variant, carriers of the rare allele are identified, and a test of the null hypothesis 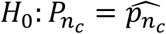 is performed testing whether *n*_*d*_ is consistent with the outcome probabilities within the carriers, based on the null model.

#### Step 1: Estimating disease probabilities under the null hypothesis

At step 1, we fit a null model under the assumption *β* = 0, using the existing penalized quasi-likelihood algorithm for logistic mixed models (15). This approach is implemented in multiple software, including the GENESIS R package (16), GMMAT (17), and SAIGE (13). In both GENESIS and GMMAT, the vector of fixed effects ***α*** and the variance component 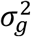 are estimated using an implementation of an AI-REML (Average Information Restricted Maximum Likelihood) algorithm on top of the penalized quasi-likelihood (PQL) approach (17), but the proposed tests do not depend on the specific algorithm used for estimating the outcome probabilities. From the fitted null model, we obtain estimates 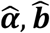, and an estimated disease probability vector by plugging them in to obtain 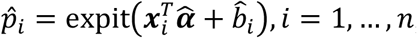, where *expit* is the inverse of the logit function. If the variance component 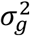 is estimated as 0, so is 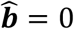, and the analysis reverts to the independent individual settings.

#### Step 2: Testing the association between a genetic variant and disease status

Suppose that we obtained disease probability estimates 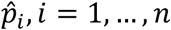, under the null as described above. Denote *n*_*c*_ as the number of carriers of the rare variant (“carriers” henceforth), i.e. those with *g* > 0, so that 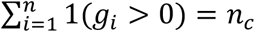. Without loss of generality, assume that participants *i* = 1, …, *n*_*c*_ are the carriers. Let *n*_*d*_ be the number of diseased carriers

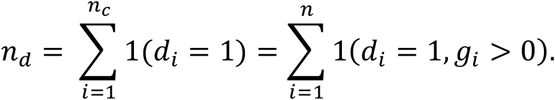

Let 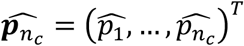 denote the vector of estimated disease probabilities for carriers of the rare variant. Despite 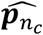 being estimated, we treat it as fixed. For testing, we assess the goodness-of-fit of the estimated model to the observed disease status in the carriers, by testing the null hypothesis:

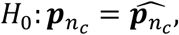

where 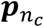 is the true, unknown, vector of outcome probabilities among the carriers.

The *p*-value for testing the null hypothesis of no variant-disease association is given by:

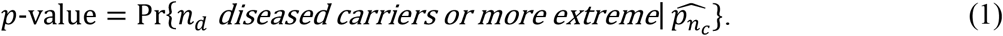

This is a two-tailed p-value, because *n*_*d*_ can appear to be lower or higher than expected. When only a single person carries the rare variant, i.e. *n*_*c*_ = 1, the calculation is trivial; Equation (1) reduces to the single carrier’s fitted probability 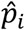 if they are a case, and 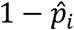 if they are a control. When *n*_*_ > 1, there are two special cases that are already developed. 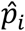 for all carriers are equal, and outcomes for all carriers are independent, then 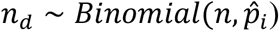, and the p-value is the tail area (possibly two tails) of the standard Binomial distribution, i.e. a Binomial exact test. If the 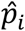 for the carriers differ but independence still holds, the distribution is the Poisson-Binomial distribution, and the test is the previously-proposed BinomiRare test for independent data (9). In the general case, an arbitrary sum of binomial variables, possibly correlated, has the Conway-Maxwell-Poisson (CMP)-Binomial distribution, which can be approximated by the CMP distribution (18; 19) when the number of carriers is “large enough” (see appendix).

In addition to the p-value above, we also study the mid-*p*-value, which was previously shown to improve properties of discrete tests (20) and to be less conservative. The mid-*p*-value is always smaller than the p-value, because when summing the tail areas probabilities, it accounts for only half of the probability of the observed event *n*_*d*_, whereas the p-value uses it as it is, without dividing in half.

### BinomiRare and CMP tests using conditional probabilities

In the appendix, we show that the distribution of *n*_*d*_ in the general case can be approximated by the CMP distribution and develop the CMP test. However, because approximations may not work well in practice for low carrier count *n*_*c*_, we also attempt a different approach. Note that for two individuals *i* and *j*, we have that *D*_*i*_ and *D*_*j*_ were independent if the true conditional disease probabilities were known. In other words, given conditional disease probabilities, knowing the disease status of individual *i* does not inform of the disease status of individual *j*. Therefore, we consider using the BinomiRare test which was developed for independent data – with the conditional probabilities. We note that this independence may not hold when probabilities are estimated, and therefore it is not trivially true that the BinomiRare is appropriate in this setting. Both the CMP and the BinomiRare tests for correlated data are available in the GENESIS R package for genetic association analysis (21).

### Simulation study: testing rare variant associations using BinomiRare and CMP in a sample of trios

We carried out a simulation study to evaluate the performance of BinomiRare and CMP tests in samples of correlated individuals. In each simulation, we generated 3,000 individuals as 1,000 trios (two parents and one offspring), as follows. For 1,000 pairs of parents, and each of two chromosomal copies, we generated 20 independent “non-causal” genetic variants by first sampling minor allele frequencies (MAF) from a uniform *U*[0.05, 0.5] distribution and setting MAF ∈ {0.05, 0.02, 0.01, 0.001} for one “causal” variant, followed by sampling of genetic variants using a Binary distribution based on these MAF. For each parent, allele count was the sum of the two sampled alleles. For each variant independently, an offspring inherited one allele from each of the parents. The parental allele was sampled at random with equal probabilities from the two alleles. We used the 21 (1 causal and 20 non-causal) simulated genotypes to generate a variable mimicking a principal component (PC), as a weighted sum of all allele counts, with weights sampled from a standard Normal distribution *N*(0,1). Next, we simulated probability of disease using a mixed logistic model:

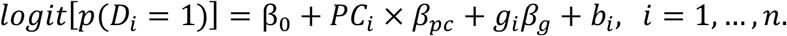

Here, exp (*β*_0_) ∈ {0.01, 0.05, 0.5} is the probability of disease in non-carriers (*g*_*i*_ = 0) with genetic PC and *b*_*i*_ equal to zero; *β*_*pc*_ models the association of the PC with disease probability, *β*_*g*_ is the effect of the (causal) variant of interest, and ***b*** = (*b*_1_, …, *b*_*n*_)^*T*^, representing the correlation across individuals, is sampled from a multivariate normal distribution 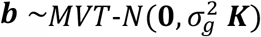, with the correlation matrix ***K*** being a block diagonal kinship matrix, having twice the kinship coefficient between a child and each of their parents, i.e. 0.5. We set 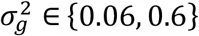. In all simulations we had *β*_.*_ = 0.1. The variant effect was varied from zero when evaluating Type 1 error rate, to *β*_)_ = log (*Odds Ratio*) ∈ {log(2), log(3), log(4)} when evaluating power. We then sampled disease status for each individual from a Binary distribution with the computed disease probability. Finally, we applied the BinomiRare and CMP tests and computed p-values and mid-p-values. We performed 1×10^7^ replicates to estimate Type 1 error rate and 1×10^5^ replicates to estimate power. We estimated Type 1 error rate and power for p-value threshold for declaring significance {1×10^−2^, 1×10^−3^, 1×10^−4^}. For tests that did not, empirically, control the Type I error rate for a given p-value threshold (i.e. the proportion of simulations passing the threshold was higher than the threshold), we computed a “calibrated threshold”, defined as a value for which the proportion of simulations with p-value less than this value was the desired threshold. We then used this calibrated threshold to estimate power, specifically power at an “honest alpha”. Our main results are those focused in simulations in which the variance component had non-zero estimate, but we analyze all simulations.

### The TOPMed whole genome sequencing Study

WGS was performed via TOPMed and the NHGRI’s Centers for Common Disease Genetics (CCDG) programs. WGS was performed using DNA from blood at multiple sequencing centers using Illumina X10 technology at an average sequencing depth of >30X. Studies and samples were sequenced in multiple phases. Periodically, the TOPMed Informatics Research Center (IRC) performed variant calling on the combined TOPMed and CCDG samples, resulting in multiple releases of data “freezes”. Details regarding sequencing methods and quality control are provided elsewhere (22) and in the TOPMed website (https://www.nhlbiwgs.org/data-sets).

We used three TOPMed multi-ethnic data sets: a data set of small vessel disease stroke (SVD stroke) in the Women Health Initiative (WHI), a study of short-sleep, and a study of venous thromboembolism (VTE), with the latter two comprised of individuals from multiple TOPMed cohorts. We performed data analysis to demonstrate the BinomiRare test. The approaches for data analysis were similar. GRMs were constructed based on the analytic datasets of each of the analyses, using all genetic variants with minor allele frequency≥0.001. Logistic mixed models under the null were fit and adjusted for age, sex, and self-reported race/ethnic group, and for short-sleep, also for parent study/cohort. SVD stroke and short sleep analyses used TOPMed freeze 5b release, while the VTE analysis used TOPMed freeze 8 genotype release. All participants provided written informed consent at their recruitment centers.

### The TOPMed WHI stroke dataset

The Women’s Health Initiative (WHI) is a long-term health study following postmenopausal women aged 50-79 years who were recruited from 1993 through 1998 from 40 clinical centers throughout the U.S. (23). In the present analysis, we focus on a subset of 5,358 WHI participants who were sequenced through TOPMed with data available via freeze 5b, and had SVD stroke case-control classification, according to the following methodology: stroke diagnosis requiring and/or occurring during hospitalization was based on the rapid onset of a neurological deficit attributable to an obstruction or rupture of an arterial vessel system. Hospitalized incident stroke events were identified by semiannual questionnaires and adjudicated following medical record review, which occurred both locally (at individual study sites) and centrally. Ischemic strokes were further classified by the central neurologist adjudicators into cardio-embolic stroke, larger artery stroke, and SVD stroke according to the Trial of Org 10172 Acute Stroke Trial (TOAST) criteria (24). The TOAST classification focuses on the presumed underlying stroke mechanism and requires detailed investigations (such as brain computed tomography, magnetic resonance imaging, angiography, carotid ultrasound, and echocardiography). Baseline stroke cases were excluded from the analysis and VTE cases were excluded from the control samples. Further, participants who had non-SVD stroke were excluded.

### The TOPMed short sleep dataset

We used sleep duration data from multiple TOPMed cohorts, as described in the Supplementary Information detailing phenotype harmonization for short sleep analysis. Short sleep was defined as self-reported sleep duration during weekday, or usual sleep (if sleep duration during the weekdays was not available) being 5 hours or less. Otherwise, if self-reported sleep duration was 6 hours or longer and less than 9 hours, sleep was “normal”. Individuals with self-reported sleep duration longer than 5 hours and shorter than 6 hours were excluded to minimize risk of misclassification. Because of a well-known “U-shaped” relationship between sleep duration and cardiovascular disease (25), suggesting that potential non-linearity in genetic associations may exist as well, we also excluded “long sleepers” reporting 9 or more hours of usual sleep,

### The TOPMed VTE dataset

The TOPMed VTE dataset includes TOPMed participants from six studies, combining prospective cohort and case-only studies. Individuals were matched across groups defined to be homogeneous with respect to race/ethnicity and sex, and strata defined by age at event (determined according to cases). The matching strategy resulted in a sample set mimicking a case-control study, with 11,627 individuals of which 3,793 are cases, and 7,834 are controls.

### Association testing of rare coding variants within known disease causing genes

For each of the SVD stroke, short sleep, and VTE datasets, we considered a known gene associated with the disorder. For stroke, we focused on the *NOTCH3* gene, in which mutations may cause Cerebral autosomal dominant arteriopathy with subcortical infarcts and leukoencephalopathy (CADASIL), which causes ischemic stroke (26). For short sleep, we focused on the gene *DEC2* (also known as *BHLHE41*), a transcription inhibitor of orexin, a neuropeptide that regulates wakefulness (27; 28). For VTE, we focused on the coagulation factor V gene, *F5* (29; 30), which has a known common variant highly associated with VTE, factor V Leiden (rs6025). We performed single variant analysis within the candidate genes, as follows. We selected a subset of rare variants within the genes based on functional annotations, with the goal of increasing power by focusing on variants that are more likely to be functional compared to others. In detail, the filter based on functional annotation included the selection of variants that were: (a) high confidence loss of function variants according to the Ensembl Variant Effect Predictor (31), (b) missense variants if they are predicted deleterious by either SIFT 4G (32), Polyphen2-HDIV (33), Polyphen2-HVAR (33), or LRT-pred (34), (c) inframe indels with FATHMM-XF coding score > 0.5 (35), or (d) variants that are synonymous according to the Ensemble Variant Effect Predictor and have FATHMM-XF coding score > 0.5. The annotation based variant filtering was performed using the Annotation explorer application on the NHLBI’s BioData Catalyst (36). We further filtered variants to those that passed the TOPMed QC filter (22), had at least 3 carriers of the rare allele, and had no more than 300 carriers. This upper threshold was defined because we were interested specifically in rare variants and because it was previously shown that properties of statistical tests of rare variant associations depend on carrier count, rather than on allele frequency (14). Finally, we further restricted the set of variants to those that had reasonable statistical power according to a power analysis performed as follows. We arbitrarily assumed an odds ratio (OR) of 2 for a causal variant, and for each variant we computed power based on a function developed for the BinomiRare test (provided here https://github.com/tamartsi/Binary_combine/blob/master/compute_power.R). The function uses the estimated outcome probabilities in the sample, an OR, the number of variant carriers, and p-value threshold, to compute power. To increase accuracy, for each variant we specifically used the estimated disease probabilities among the variant carriers.

## Results

### Simulations studies

We studied the performance of the tests in simulations of 1,000 trios. In the setting where 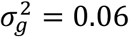, about half of the simulations estimated the variance component to be zero. When 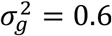, this happened in about a third of the simulations. The number of carriers of the simulated rare variant allele was in the range [0, 27] when MAF = 0.001, [17, 119] when MAF = 0.01, [58, 203] when MAF = 0.02, and [195, 401] when MAF = 0.05. Table 1 provides estimated Type 1 error rates in the simulations, restricted to those simulations in which the estimated variance component was 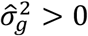. For BinomiRare, we only provide results for the mid-*p*-value, because in our simulations it always controlled the Type 1 error rate, while the usual p-value controlled it as well while being more conservative. For CMP, we only provide results for the usual p-value, because it sometimes did not control the type 1 error and the lack of control was worse with the mid-*p*-value. The CMP test usually did not control the type 1 error when the variant was very rare (MAF =0.001), and when the case proportion was low (exp(*β*_0_) = 0.05). Its performance improved as the MAF increased. In the Supplementary Information, Table S3-S5, we provide complete simulation results, including both mid-*p*-value and the usual p-value for both the CMP and BinomiRare tests, and results computed over all simulations, and computed over the simulations in which 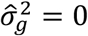.

**Table 1:**
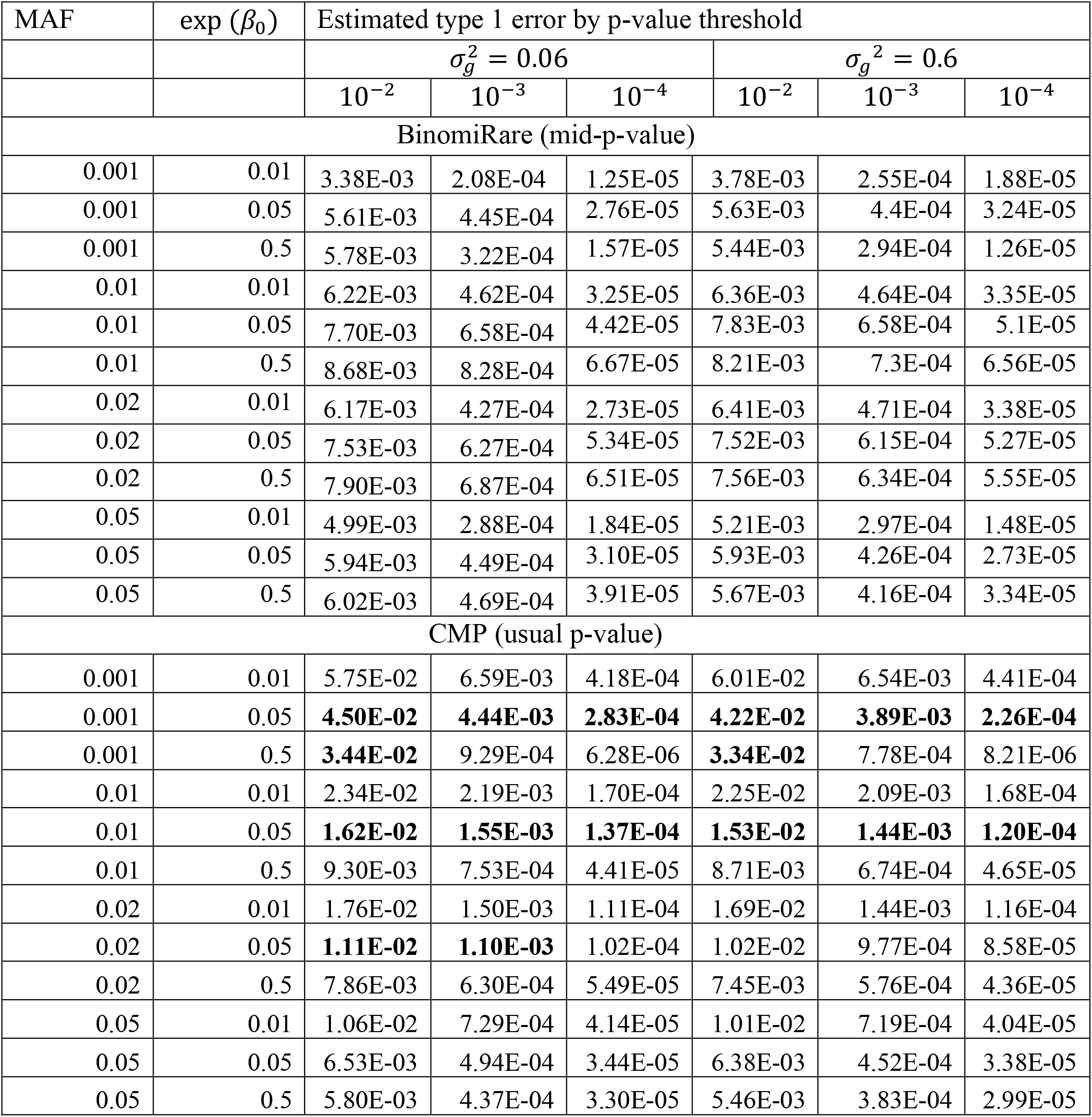
Estimated Type 1 error rates of BinomiRare and CMP tests in simulations with related individuals. Bolded numbers highlight settings in which the Type 1 error was not controlled, defined according to Type 1 error rate being larger than the highest value in a 95% confidence intervals around the expected Type 1 error rate, based on Binomial distribution with parameters being the p-value threshold and number of simulations used.

| MAF | exp ($\beta_0$) | Estimated type 1 error by p-value threshold | | | | | |
| --- | --- | --- | --- | --- | --- | --- | --- |
| | | $\sigma_g^2 = 0.06$ | | | $\sigma_g^2 = 0.6$ | | |
| | | $10^{-2}$ | $10^{-3}$ | $10^{-4}$ | $10^{-2}$ | $10^{-3}$ | $10^{-4}$ |
| BinomiRare (mid-p-value) |  |  |  |  |  |  |  |
| 0.001 | 0.01 | 3.38E-03 | 2.08E-04 | 1.25E-05 | 3.78E-03 | 2.55E-04 | 1.88E-05 |
| 0.001 | 0.05 | 5.61E-03 | 4.45E-04 | 2.76E-05 | 5.63E-03 | 4.4E-04 | 3.24E-05 |
| 0.001 | 0.5 | 5.78E-03 | 3.22E-04 | 1.57E-05 | 5.44E-03 | 2.94E-04 | 1.26E-05 |
| 0.01 | 0.01 | 6.22E-03 | 4.62E-04 | 3.25E-05 | 6.36E-03 | 4.64E-04 | 3.35E-05 |
| 0.01 | 0.05 | 7.70E-03 | 6.58E-04 | 4.42E-05 | 7.83E-03 | 6.58E-04 | 5.1E-05 |
| 0.01 | 0.5 | 8.68E-03 | 8.28E-04 | 6.67E-05 | 8.21E-03 | 7.3E-04 | 6.56E-05 |
| 0.02 | 0.01 | 6.17E-03 | 4.27E-04 | 2.73E-05 | 6.41E-03 | 4.71E-04 | 3.38E-05 |
| 0.02 | 0.05 | 7.53E-03 | 6.27E-04 | 5.34E-05 | 7.52E-03 | 6.15E-04 | 5.27E-05 |
| 0.02 | 0.5 | 7.90E-03 | 6.87E-04 | 6.51E-05 | 7.56E-03 | 6.34E-04 | 5.55E-05 |
| 0.05 | 0.01 | 4.99E-03 | 2.88E-04 | 1.84E-05 | 5.21E-03 | 2.97E-04 | 1.48E-05 |
| 0.05 | 0.05 | 5.94E-03 | 4.49E-04 | 3.10E-05 | 5.93E-03 | 4.26E-04 | 2.73E-05 |
| 0.05 | 0.5 | 6.02E-03 | 4.69E-04 | 3.91E-05 | 5.67E-03 | 4.16E-04 | 3.34E-05 |
| CMP (usual p-value) |  |  |  |  |  |  |  |
| 0.001 | 0.01 | 5.75E-02 | 6.59E-03 | 4.18E-04 | 6.01E-02 | 6.54E-03 | 4.41E-04 |
| 0.001 | 0.05 | **4.50E-02** | **4.44E-03** | **2.83E-04** | **4.22E-02** | **3.89E-03** | **2.26E-04** |
| 0.001 | 0.5 | **3.44E-02** | 9.29E-04 | 6.28E-06 | **3.34E-02** | 7.78E-04 | 8.21E-06 |
| 0.01 | 0.01 | 2.34E-02 | 2.19E-03 | 1.70E-04 | 2.25E-02 | 2.09E-03 | 1.68E-04 |
| 0.01 | 0.05 | **1.62E-02** | **1.55E-03** | **1.37E-04** | **1.53E-02** | **1.44E-03** | **1.20E-04** |
| 0.01 | 0.5 | 9.30E-03 | 7.53E-04 | 4.41E-05 | 8.71E-03 | 6.74E-04 | 4.65E-05 |
| 0.02 | 0.01 | 1.76E-02 | 1.50E-03 | 1.11E-04 | 1.69E-02 | 1.44E-03 | 1.16E-04 |
| 0.02 | 0.05 | **1.11E-02** | **1.10E-03** | 1.02E-04 | 1.02E-02 | 9.77E-04 | 8.58E-05 |
| 0.02 | 0.5 | 7.86E-03 | 6.30E-04 | 5.49E-05 | 7.45E-03 | 5.76E-04 | 4.36E-05 |
| 0.05 | 0.01 | 1.06E-02 | 7.29E-04 | 4.14E-05 | 1.01E-02 | 7.19E-04 | 4.04E-05 |
| 0.05 | 0.05 | 6.53E-03 | 4.94E-04 | 3.44E-05 | 6.38E-03 | 4.52E-04 | 3.38E-05 |
| 0.05 | 0.5 | 5.80E-03 | 4.37E-04 | 3.30E-05 | 5.46E-03 | 3.83E-04 | 2.99E-05 |

In power analysis, after appropriately calibrating the p-value threshold for the CMP test, CMP was either equally powerful as BinomiRare or more powerful (see Figures S1-S3 in the Supplementary Information). The patterns were similar across p-value thresholds used, and across the two variance component parameters used in the simulations. Notably, when the disease was common (exp (*β*_0_) = 0.5) the power was lower when the variance component was high 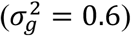 compared to when it was low 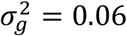. When the disease was rare (exp(*β*_0_ = 0.05), the power was essentially the same with both values of variance components.

### Data analysis: TOPMed data sets

For each of the three TOPMed datasets that we considered, Table 2 provides the sample sizes, gene of interest, and number of variants according to sequential filtering: the number of available (non-monomorphic) variants in the sample that passed the functional filters described in the Methods section, number of variants after applying quality filters, and after restricting to those with at least 3 carriers of the rare alleles and less than 300 carriers, and the number of variants with at least 50% power to reject the null hypothesis at the 0.05 level under the assumption of odds ratio =2. There were 3 such variants in the *NOTCH3*-SVD stroke analysis, 1 variant in the *DEC2*-short sleep analysis, and 4 variants in the *F5*-VTE analysis.

**Table 2:** Characteristics of the TOPMed datasets and variants considered for association testing.

|  | SVD stroke | Short sleep | VTE |
| --- | --- | --- | --- |
| # individuals in the analysis | 5,358 | 20,021 | 11,627 |
| # cases | 692 (12.9%) | 2,408 (12%) | 3,793 (32.6%) |
| # controls | 4,666 (87.1%) | 17,613 (88%) | 7,834 (67.4%) |
| Gene of interest | *NOTCH3* | *DEC2/*<br>*BHLHE41* | *F5* |
| # potentially functional non-monomorphic variants identified | 122 | 58 | 142 |
| # variants further passing TOPMed quality filters | 117 | 49 | 132 |
| # variants further having 2<carriers <300 | 20 | 9 | 25 |
| # variants with estimated power >0.5 at the 0.05 $\alpha$ level | 3 | 1 | 4 |

Table 3 provides the results from testing each of the variants passing this estimated power filter. Of the three tested *NOTCH3* variants, rs115582213 had p-value=0.03. For short sleep, only a single *DC2* variant was tested, it had BinomiRare mid-p-value=0.03, suggesting association with short sleep. For the *F5* gene and VTE, none of the four tested variants showed evidence of association.

**Table 3:** Results from association analysis of rare genetic variants within monogenic disease genes of interest. Genetic variants presented are those that passed functional annotation and statistical power filters. For each variant we provide its BinomiRare p-value and mid-p-value, the number of carriers of the rare allele *n*_*c*_, the number of carriers with the outcome *n*_*d*_, the estimated power computed while assuming effect size OR=2 and p-value threshold=0.05, pathogenicity interpretation from ClinVar, CADD score, and FATHMM-XF coding score.

| rsID | Variant | BinomiRare<br>pval | BinomiRare<br>midp | $n_c$ | $n_d$ | Estimated<br>Power<br>(OR=2) | ClinVar<br>Interpretation | CADD<br>PHRED | FATHMM-XF coding |
| --- | --- | --- | --- | --- | --- | --- | --- | --- | --- |
| SVD Stroke: *NOTCH 3* gene |  |  |  |  |  |  |  |  |  |
| rs115582213 | chr-19-15162524-C-T | 0.04 | 0.03 | 87 | 17 | 0.7 | Benign/<br>Likely<br>benign | 25.4 | 0.66 |
| rs112197217 | chr-19-15179425-G-T | 0.53 | 0.49 | 166 | 23 | 0.91 | Benign/<br>Likely benign | 21 | 0.42 |
| rs11670799 | chr-19-15188240-G-A | 0.81 | 0.77 | 180 | 23 | 0.94 | Benign/<br>Likely benign | 28.8 | 0.68 |
| Short sleep: *DEC2* gene |  |  |  |  |  |  |  |  |  |
| rs121912617 | chr-12-26122364-G-T | 0.04 | 0.03 | 127 | 38 | 0.98 | Not available | 27.5 | 0.66 |
| VTE: *F5* gene |  |  |  |  |  |  |  |  |  |
| rs6026 | chr-1-169528054-C-T | 0.37 | 0.34 | 115 | 31 | 0.94 | Benign/<br>Likely benign | 25.7 | 0.75 |
| rs6034 | chr-1-169529782-G-C | 1.00 | 0.94 | 46 | 16 | 0.57 | Conflicting<br>interpretations | 21.3 | 0.56 |
| rs78958618 | chr-1-169542985-G-A | 0.67 | 0.63 | 130 | 32 | 0.94 | Benign | 15.18 | 0.11 |
| rs9332485 | chr-1-169586344-C-T | 0.37 | 0.34 | 222 | 55 | 1 | Benign/<br>Likely benign | 22.5 | 0.23 |

## Discussion

We extended the BinomiRare test and proposed a CMP test for testing the association of a rare genetic variant with a binary outcome in the mixed model framework. These tests were specifically developed to handle variants with very low minor allele counts (tens of carriers), because it was previously shown that other tests that allow for covariates adjustment such as the naïve Score test and the SPA test do not always control the type 1 error in the very low count settings (14). Both carriers-only tests first estimate the outcome probabilities for each person in a data set, while accounting for covariates and for genetic relatedness (and possibly other covariance matrices) via a mixed model, and then use the estimated conditional disease probabilities. For a single variant, the carriers of the rare alleles are identified, and based on their disease probabilities and the observed number of “cases”, a p-value is computed, as the probability of observing the given number of cases or more extreme given the estimated outcome probabilities. The BinomiRare test with conditional probabilities performed well, while, surprisingly, the CMP test did not control the Type 1 error rate for settings with low carrier counts. This was likely because the approximations on which it relies are asymptotic in its non-centrality parameter *λ*, which is related to the number of carriers.

We demonstrated the application of the BinomiRare test using three TOPMed studies: of SVD stroke, short sleep, and VTE. Due to the low power for testing low-count variants, we filtered variants according to functional annotation, and according to computed statistical power. The limitation of this approach is that (1) The deleteriousness predicting annotations used and the filters applied to them may not have captured the true functional variant set; (2) the power analysis was based on an arbitrarily selected OR parameters. In this study, we chose OR=2 and only considered the handful of variants that had estimated power > 0.5 for testing while requiring p-value (*α* level)<0.05. We recognize that many rare variants have larger effect sizes. However, if we specified a larger OR parameter, and thus included more variants in our analysis, a more stringent *α* level would be needed. Thus, the resulting list of variants to test may have been similar. More work is needed developing strategies for identifying single rare variant associations.

For each of the phenotypes, SVD stroke, VTE, and short sleep, we searched for rare variants within genes with known trait associations. For SVD stroke, we considered *NOTCH3*, because some *NOTCH3* variants have been in CADASIL patients, which poses a risk for stroke. Most *NOTCH3* mutations reported as associated with CADASIL are those involving loss or gain of a cysteine residue, leading to unpaired cysteine (37). Single nucleotide variants in *NOTCH3* have not yet consistently identified as associated with SVD stroke in population-based studies. Here, we identified the rare variant rs115582213 (BinomiRare mid-p-value=0.03). This variant was rare, with 87 variant carriers out of 5,358 individuals in the dataset. Of these, 17 individuals had SVD stroke.

For VTE, we considered the *F5* gene. The *F5* gene harbors the strongest known, relatively common, genetic risk factor for VTE, the rs6025 variant (38; 39). This motivated the search for rare variants in this gene. We did not identify any variant associated with VTE at the p-value<0.05 level. We did not consider rs6025 as part of our testing strategy because it was common with MAF=0.04, and had 839 carriers of the rare allele, a setting in which other tests such as the SPA should be able to control the type 1 error well and also be more powerful. Still, as a positive control we tested its association with VTE using BinomiRare, and the p-value was 1.5×10^−14^.

Short sleep has been consistently associated with cardiovascular and cardiometabolic disease (40; 41). Genetic determinant of short sleep may help elucidate this connection (42). We considered the *DEC2/BHLHE4* gene, which a mutation with a know familial aggregation associated with short sleep. Our filtering strategy resulted in a single variant considered for testing: rs121912617, the known short sleep mutation (27). In our data, it was associated with short sleep with BinomiRare mid-p-value=0.03. Rs121912617 is substantially more common (yet is still rare) in African Americans compared to European Americans (0.01 MAF in African Americans from the TOPMed short sleep datasets, compared to MAF < 0.001 in European Americans from the same dataset), allowing for observing this association in a population-based, rather than a family-based, study.

Here, we demonstrated the BinomiRare test for testing single-variant associations in data with known or cryptic relatedness. It can also be used to test sets of rare variants, by defining a carrier as an individual with at least one rare allele in the variant set. It is a topic of future research to extend this framework to use the counts of the rare variant allele and increase power.

## Data Availability

This manuscript used genome-sequencing data from TOPMed, and phenotypes from TOPMed participating studies. These data are available by application to dbGaP using study-specific accessions (phs numbers). WGS for "NHLBI TOPMed: Genetics of Cardiometabolic Health in the Amish" (phs000956) was performed at the Broad Institute of MIT and Harvard (3R01HL121007-01S1). WGS for "NHLBI TOPMed: Cleveland Family Study - WGS Collaboration" (phs000954) was performed at the University of Washington Northwest Genomics Center (3R01HL098433-05S1, HHSN268201600032I). WGS for "NHLBI TOPMed: Cardiovascular Health Study" (phs001368.v2.p2) was performed at Baylor Genome Sequencing Center (3U54HG003273-12S2, HHSN268201500015C, HHSN268201600033I). WGS for "NHLBI TOPMed: Framingham Heart Study" (phs000974) was performed at the Broad Institute of MIT and Harvard (3R01HL092577-06S1, 3U54HG003067-12S2). WGS for "NHLBI TOPMed: Heart and Vascular Health Study (HVH)" (phs000993.v4.p2) was performed at Baylor Genome Sequencing Center (3U54HG003273-12S2, HHSN268201500015C). WGS for "NHLBI TOPMed: Jackson Heart Study" (phs000964) was performed at the University of Washington Northwest Genomics Center (HHSN268201100037C). WGS for "NHLBI TOPMed: Multi Ethnic Study of Atherosclerosis" (phs001416) was performed at the Broad Institute of MIT and Harvard (3R01HL092577-06S1, 3U54HG003067-12S2). WGS for "NHLBI TOPMed: Whole Genome Sequencing of Venous Thromboembolism (WGS or VTE)" (phs001402.v2.p1) was performed at Baylor Genome Sequencing Center (3U54HG003273-12S2, HHSN268201500015C). WGS for "NHLBI TOPMed: Women's Health Initiative (WHI)" (phs001237.v2.p1) was performed at the Broad Institute of MIT and Harvard (HHSN268201500014C).

## Acknowledgements

This work was supported by the National Heart Lung and Blood Institute grant R35HL135818. Study-specific acknowledgements are available in the Supplementary Information. Whole genome sequencing (WGS) for the Trans-Omics in Precision Medicine (TOPMed) program was supported by the National Heart, Lung and Blood Institute (NHLBI). See Supplementary Information for sequencing center support information. Centralized read mapping and genotype calling, along with variant quality metrics and filtering were provided by the TOPMed Informatics Research Center (3R01HL-117626-02S1; contract HHSN268201800002I).

Phenotype harmonization, data management, sample-identity QC, and general study coordination were provided by the TOPMed Data Coordinating Center (R01HL-120393; U01HL-120393; contract HHSN268201800001I). We gratefully acknowledge the studies and participants who provided biological samples and data for TOPMed. The views expressed in this manuscript are those of the authors and do not necessarily represent the views of the National Heart, Lung, and Blood Institute; the National Institutes of Health; or the U.S. Department of Health and Human Services.

## Author contributions

T.S. and E.S. developed the BinomiRare and CMP tests for correlated individuals. T.S. performed simulation studies. T.S., J.L. N.K., applied the BinomiRare test on the stroke, short sleep, and VTE datasets. J.L. Harmonized the short sleep phenotype. D.J. and B.H. used the variant annotations to define filters of likely functional variants. T.S., S.M.G., and M.P.C. implemented the BinomiRare and CMP tests in the GENESIS R package. T.S., C.A.L., N.P., N.L.S., B.J.C., A.S., and K.R., developed and implemented the strategy for constructing the VTE dataset. Y.H. harmonized the stroke phenotypes. T.S., Y.H., C.K. J.H., A.J., and A.P.R., developed the approach for analyzing the SVD stroke dataset. T.S., J.L., D.G., S.R., developed the approach for constructing the short sleep dataset. H.C., J.O., M.Z., and S.G. contributed to short sleep phenotype harmonization and discussion about analyses. R.S.V., A.D.J., A.C., A.C.M. E.B., B.P., W.T.L., J.I.R., K.D.T., S.R., X.G., M.C., C.J., S.S.R. helped collect and distribute phenotypic and genetic data and design cohort participations in TOPMed.

## APPENDIX

### The CMP test

Let 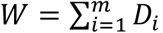, for *D*_*i*_ ∼ *Binom*(*p*_*i*_, 1) be a random variable with the CMP-Binomial probability function. When *m* increases, this distribution is approximated by the CMP distribution (Theorem 4.1. in Daly and Gaunt (43)) so that *W* ∼ *CMP*(*λ, v*). Consider proposition 2 in Kadane (19) stating:

#### Proposition 2 (Kadane, 2016)

*Suppose D*_1_, …, *D*_*m*_ *take values on* {0,1}. *Let* 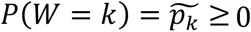, *where* 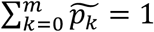. *Then there exists a unique distribution on D*_1_, …, *D*_*m*_ *such that D*_1_, …, *D*_*m*_ *are exchangeable of order m, and* 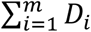 *has the same distribution as W*.

According to this proposition, an arbitrary sum of binary variables is distributed as a sum of exchangeable binary variables, where the exchangeable variables are such that there is a unique combination of probability parameter *p* = pr(*D*_1_ = 1) = ⋯ = Pr(*D*_4_ = 1) and a parameter *ρ* modeling the dependency between each pair *D*_*i*_, *D*_*j*_, *i* ≠ *j*. Therefore, two parameters suffice to characterize the distribution of an arbitrary sum of binary variables. Specifically, for a given set of carriers of a rare genetic variant, the sum of their disease statuses

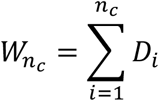

is distributed like a unique sum of exchangeable binary variables. Based on the estimated disease probabilities, we estimate the two parameters (different than the probability and dependency parameters *p* and *ρ* above) of the CMP distribution to obtain an estimated probability function in a variation of a method-of-moment approach that is based on estimated probabilities, rather than on the observed data. Daly and Gaunt (43) provided an approximation to the CMP distribution:

#### Proposition 2.3. (Daly and Gaunt)

*Let W* ∼ *CMP*(*λ, v*). *Then, for k* ∈ ℕ,

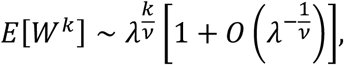

*as λ*→ ∞.

Assuming that *λ*^−1/*v*^ is small (which, as we shall see, is true when *λ* is very large, because *v* tends to be well bounded), we get that, approximately:

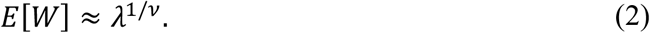

Daly and Gaunt also showed, in their equation 2.4 and based on the result in Shmueli et al (18), that:

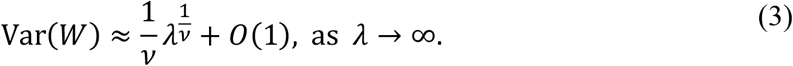

Therefore, noting that Var(*W*) = *E*[*W*^2^] − (*E*[*W*])^2^, once we estimate *E*[*W*] and *E*[*W*^2^], we use (4) and (5) to obtain estimators of *λ* and *v* by:

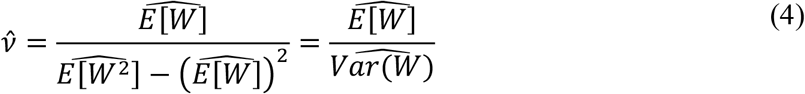

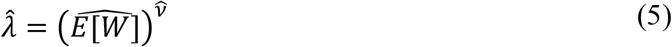

#### Estimating parameters of the CMP distribution from estimated diseased probabilities

We consider two approaches to estimate components of 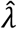 and 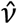, i.e. *E*[*W*], *E*[*W*^2^], and Var(*W*): an analytic approach, and a sampling-based approach. In the analytic approach, we compute 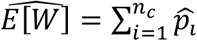, and 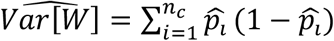. In the sampling-based approach, we generate random variables 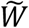 with the same distribution as 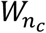 (the sum of disease statuses among the *n*_*c*_ carriers of a genetic variant), and treat them as observed data to estimate the desired quantities. More specifically, let 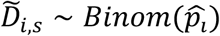 be the sampled disease status of the *i*th individual in the *s* = 1, …, *S* sample. Then:

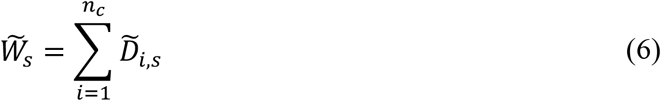

and we estimate:

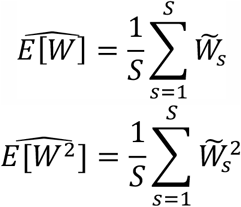

To summarize, to calculate the p-value and the mid-*p*-value, formally given by:

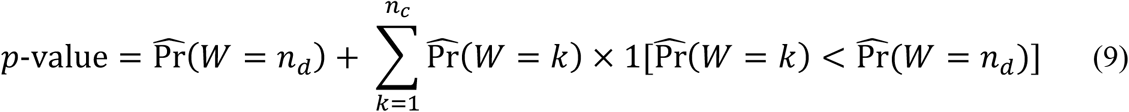

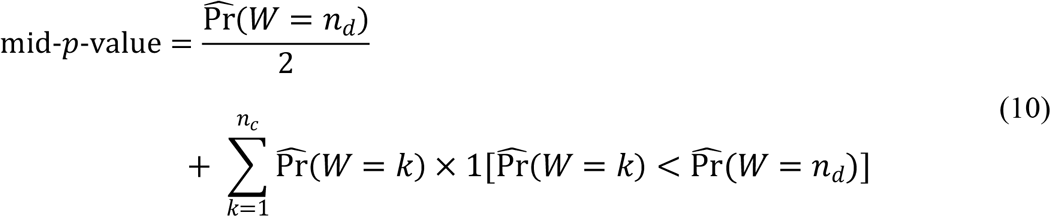

we estimate probabilities for each potential number of disease carriers, in the following process:

1. Obtain individual disease probability estimates 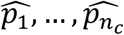 via standard approaches (e.g. logistic mixed model).
2. Compute estimates 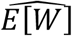 and 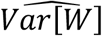 in the analytic approach, or compute 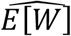 and 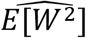 in the sampling approach.
3. Compute estimates 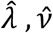 using (6) and (7).
4. Compute 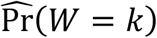 for *k* = 1, …, *n*_c_ using the R package COMPoissonReg (44).

## Notes

### Competing Interest Statement

The authors have declared no competing interest.

### Funding Statement

This work was supported by the National Heart Lung and Blood Institute grant R35HL135818. Whole genome sequencing (WGS) for the Trans-Omics in Precision Medicine (TOPMed) program was supported by the National Heart, Lung and Blood Institute (NHLBI). See Supplementary Note 2 for sequencing center support information. Centralized read mapping and genotype calling, along with variant quality metrics and filtering were provided by the TOPMed Informatics Research Center (3R01HL-117626-02S1; contract HHSN268201800002I). Phenotype harmonization, data management, sample-identity QC, and general study coordination were provided by the TOPMed Data Coordinating Center (R01HL-120393; U01HL-120393; contract HHSN268201800001I). 

### Author Declarations

Mass General Brigham IRB

