## Supplementary Information for "BinomiRare: A carriers-only test for association of rare genetic variants with a binary outcome for mixed models and any case-control proportion"

Sofer T, et al.

### 1. Phenotype harmonization for short sleep analysis

We first harmonized a sleep duration phenotype, followed by a definition of a short sleep variables. For sleep duration measures in each study, Table S1 provides the specific question or measure, the number of participants with non-missing values for this outcome, and the number of participants with non-missing values for this outcome after excluding extreme values (sleep duration < 2 hours or >12 hours), and the number of individuals after further excluding individuals from race/ethnic groups with small sample size within the cohort (e.g. we excluded 3 Hispanic Americans (HA) from FHS, because of the low number of HAs in FHS), or from ill-defined race/ethnic groups (e.g. "other"). Table S2 provides sample sizes after prioritization of questions regarding sleep duration. For example, for MESA, we prioritized self-reported sleep duration from visit 5. Thus, if an individual from MESA was included based on their self-reported sleep duration at visit 5, this individual was not included in the row referring to MESA visit 4. Rather, the table row is corresponding to MESA visit 4 only when these individuals who did not report sleep duration at visit 5. There were 22,398 individuals with a valid sleep duration phenotype.

Next, we defined short sleep variables. We considered short sleep as a sleep duration ≤5 hours, and long sleep as sleep duration ≥9 hours. For analysis of short sleep, we removed long sleepers, and also individuals reporting sleep duration “between 5-6 hours”, because these cannot be determined to be either short or normal sleepers, according to our definitions. This resulted in 20,021 individuals, 3,793 short sleepers, and 7,834 controls.

**Table S1:** Studies with sleep duration related measures/questions.

| Study | Exam/ questionnaire | Question/measure (code name in data base) | Recoded name | N | N after exclusion of extreme values | N after exclusion of small count and ill-defined backgrounds |
|---|---|---|---|---|---|---|
| Amish | HAPI Diary | wd.dur | sleep_duration_weekday | 202 | 202 | 202 |
| Amish | Longevity Diary | wd.dur | sleep_duration_weekday | 81 | 81 | 81 |
| ARIC | Exam 5 / RSE | rse21 | usual_sleep_hours | 826 | 826 | 826 |
| ARIC | SHHS exam 1 | hrswd02 | sleep_duration_weekday | 477 | 477 | 477 |
| ARIC | SHHS exam 1 | tfawdh02/tfawdm02/tfawda02/ twuwdh02/twuwdm02/twuwda02* | sleep_duration_weekday_calculated | 466 | 463 | 463 |
| CFS | Exam 2 | dayhrs/daymin* | sleep_duration_weekday | 451 | 451 | 451 |
| CFS | Exam 3 | dayhrs/daymin* | sleep_duration_weekday | 69 | 69 | 69 |
| CFS | Exam 5 | dayhrs/daymin* | sleep_duration_weekday | 580 | 578 | 578 |
| CFS | Exam 1 | daybed/endbed* | sleep_duration_weekday_calculated | 424 | 412 | 412 |
| CFS | Exam 2 | daybed/endbed* | sleep_duration_weekday_calculated | 450 | 440 | 440 |
| CFS | Exam 3 | daybed/endbed* | sleep_duration_weekday_calculated | 69 | 68 | 68 |
| CFS | Exam 4 | daybed/endbed* | sleep_duration_weekday_calculated | 206 | 197 | 197 |
| CFS | Exam 5 | daybed/endbed* | sleep_duration_weekday_calculated | 579 | 561 | 561 |
| CHS | SHHS exam 1 | hrswd02 | sleep_duration_weekday | 37 | 37 | 32 |
| CHS | SHHS exam 1 | tfawdh02/tfawdm02/tfawda02/ twuwdh02/twuwdm02/twuwda02* | sleep_duration_weekday_calculated | 36 | 36 | 31 |
| FHS | FHS- exam 1_7s | g689 | usual_sleep_hours | 1741 | 1739 | 1663 |
| FHS | FHS- exam 1-8s | h480 | usual_sleep_hours | 1578 | 1573 | 1551 |
| FHS | FHS- exam 1-8s | h726 | sleep_duration_weekday | 1551 | 1550 | 1532 |
| FHS | FHS- exam 1_9s | j628 | usual_sleep_duration | 1262 | 1256 | 1231 |

| | | | | | | |
|---|---|---|---|---|---|---|
| FHS | FHS- exam 1_9s | j921/j922* | actual_sleep_duration | 1239 | 1237 | 1212 |
| FHS | FHS- exam 1_9s | j913/j914/j915/j918/j919/j920* | usual_sleep_duration_calculated | 1221 | 1171 | 1149 |
| FHS | SHHS | hrswd02 | sleep_duration_weekday | 1472 | 1471 | 1455 |
| FHS | SHHS | tfawdh02/tfawdm02/tfawda02/ twuwdh02/twuwdm02/twuwda02* | sleep_duration_weekday_calculated | 1491 | 1483 | 1468 |
| FHS-gen3 | gen3 exam 1 | g3a596 | usual_sleep_hours | 1338 | 1336 | 1309 |
| JHS | Sleep ancillary study | weekday_sleep_duration | sleep_duration_weekday_calculated | 572 | 567 | 567 |
| | Sleep ancillary study | hrs_sleep_weekdays | sleep_duration_weekday | 573 | 572 | 572 |
| | JHS exam 1 | mhxa7 | usual_sleep_hours | 3047 | 3041 | 3041 |
| | JHS exam 3 | slea1 | sleep_duration_weekday | 2272 | 2261 | 2261 |
| MESA | Exam 4 | slpwkhr4 | sleep_duration_weekday | 3991 | 3987 | 3987 |
| | Exam 5 | wkdaysleepdur5t | sleep_duration_weekday | 1815 | 1799 | 1799 |
| | Exam 5 | bedtmwkday5c/waketmwkday5c* | sleep_duration_weekday_calculated | 1811 | 1790 | 1790 |
| WHI | Form f37 | hrsslp** | usual_sleep_hours | 10006 | 10006 | 9931 |

**Table S2:** Studies and sleep duration measures prioritization. For each study, we provide the sample size from each visit and question that were selected, where the first row of each study represents the question/visit with highest priority, so that all individuals who responded to this question in this visit and had appropriate values were selected. The second row in a study represent the next question/visit in the priority order for that study. N represent the number of people with valid responses for that question/visit, and that did not have a valid response in the higher priority question, and so on. For each study, we also provide the total sample size, representing all people who responded with valid replies to at least one of the questions/visits.

| Study | Exam/questionnaire | Question/measure (by priority) | N | Study total |
|---|---|---|---|---|
| Amish | HAOI-diary | sleep_duration_weekday | 202 | |
| Amish | Longevity-diary | sleep_duration_weekday | 74 | 276 |
| ARIC | SHHS exam 1 | sleep_duration_weekday | 477 | |
| ARIC | SHHS exam 1 | sleep_duration_weekday_calculated | 2 | |
| ARIC | Exam 5 | usual_sleep_hours | 594 | 1073 |
| CFS | Exam 5 | sleep_duration_weekday | 578 | |
| CFS | Exam 5 | sleep_duration_weekday_calculated | 2 | |
| CFS | Exam 4 | sleep_duration_weekday_calculated | 68 | |
| CFS | Exam 3 | sleep_duration_weekday | 16 | |
| CFS | Exam 2 | sleep_duration_weekday | 169 | |
| CFS | Exam 1 | sleep_duration_weekday_calculated | 32 | 865 |
| CHS | SHHS exam 1 | sleep_duration_weekday | 32 | 32 |
| FHS | SHHS exam 1 | sleep_duration_weekday | 1455 | |
| FHS | SHHS exam 1 | sleep_duration_weekday_calculated | 35 | |
| FHS | Exam 1-8s | usual_sleep_hours | 308 | |
| FHS | Exam 1-8s | sleep_duration_weekday | 2 | |
| FHS | Exam1-7s | usual_sleep_hours | 2 | 1802 |
| FHS-gen3 | gen3- Exam 1 | usual_sleep_hours | 1309 | 1309 |
| JHS | Sleep ancillary study | sleep_duration_weekday | 572 | |
| JHS | Sleep ancillary study | sleep_duration_weekday_calculated | 2 | |
| JHS | Exam 3 | sleep_duration_weekday | 1698 | |
| JHS | Exam 1 | usual_sleep_hours | 824 | 3096 |
| MESA | Exam 5 | sleep_duration_weekday | 1799 | |
| MESA | Exam 4 | sleep_duration_weekday | 2215 | 4014 |
| WHI | From 37 | usual_sleep_hours | 9931 | 9931 |
| | | | | 22398 |

### 2. Type 1 error rate estimates from simulations.

Table S3: estimated Type 1 error rates in simulations for each of the BinomiRare and CMP tests, in each of the settings. The Type 1 error rates here were estimated using simulations in which the variance component was estimated as non-zero. Orange colored cells correspond to settings in which the Type 1 error rates were higher than desired.

| Test | MAF | Odds | Mid-*p*-value | | | p-value | | |
|---|---|---|---|---|---|---|---|---|
| | | | Threshold: 1E-02 | 1E-03 | 1E-04 | Threshold: 1E-02 | 1E-03 | d1E-04 |
| | | | $\sigma_g^2 = 0.6$ | | | | | |
| | 0.001 | 0.01 | 3.78E-03 | 2.55E-04 | 1.88E-05 | 2.19E-03 | 1.22E-04 | 3.78E-03 |
| BinomiRare | 0.001 | 0.05 | 5.63E-03 | 4.40E-04 | 3.24E-05 | 2.81E-03 | 2.15E-04 | 1.74E-05 |
| | 0.001 | 0.5 | 5.44E-03 | 2.94E-04 | 1.26E-05 | 2.58E-03 | 1.28E-04 | 5.40E-06 |
| | 0.01 | 0.01 | 6.36E-03 | 4.64E-04 | 3.35E-05 | 3.49E-03 | 2.43E-04 | 1.80E-05 |
| | 0.01 | 0.05 | 7.83E-03 | 6.58E-04 | 5.10E-05 | 5.05E-03 | 3.98E-04 | 2.82E-05 |
| | 0.01 | 0.5 | 8.21E-03 | 7.30E-04 | 6.56E-05 | 6.71E-03 | 5.69E-04 | 4.74E-05 |
| | 0.02 | 0.01 | 6.41E-03 | 4.71E-04 | 3.38E-05 | 3.65E-03 | 2.53E-04 | 1.64E-05 |
| | 0.02 | 0.05 | 7.52E-03 | 6.15E-04 | 5.27E-05 | 5.46E-03 | 4.19E-04 | 3.28E-05 |
| | 0.02 | 0.5 | 7.56E-03 | 6.34E-04 | 5.55E-05 | 6.53E-03 | 5.29E-04 | 4.49E-05 |
| | 0.05 | 0.01 | 5.21E-03 | 2.97E-04 | 1.48E-05 | 3.30E-03 | 1.71E-04 | 7.89E-06 |
| | 0.05 | 0.05 | 5.93E-03 | 4.26E-04 | 2.73E-05 | 4.81E-03 | 3.35E-04 | 2.08E-05 |
| | 0.05 | 0.5 | 5.67E-03 | 4.16E-04 | 3.34E-05 | 5.14E-03 | 3.66E-04 | 2.93E-05 |
| | 0.001 | 0.01 | 6.83E-02 | 1.38E-02 | 1.05E-03 | 6.01E-02 | 6.54E-03 | 4.41E-04 |
| CMP | 0.001 | 0.05 | 7.39E-02 | 8.19E-03 | 5.13E-04 | 4.22E-02 | 3.89E-03 | 2.26E-04 |
| | 0.001 | 0.5 | 6.70E-02 | 2.51E-03 | 3.35E-05 | 3.34E-02 | 7.78E-04 | 8.21E-06 |
| | 0.01 | 0.01 | 3.83E-02 | 3.84E-03 | 3.21E-04 | 2.25E-02 | 2.09E-03 | 1.68E-04 |
| | 0.01 | 0.05 | 2.27E-02 | 2.32E-03 | 2.04E-04 | 1.53E-02 | 1.44E-03 | 1.20E-04 |
| | 0.01 | 0.5 | 1.07E-02 | 8.71E-04 | 6.25E-05 | 8.71E-03 | 6.74E-04 | 4.65E-05 |
| | 0.02 | 0.01 | 2.75E-02 | 2.51E-03 | 2.15E-04 | 1.69E-02 | 1.44E-03 | 1.16E-04 |
| | 0.02 | 0.05 | 1.35E-02 | 1.40E-03 | 1.34E-04 | 1.02E-02 | 9.77E-04 | 8.58E-05 |
| | 0.02 | 0.5 | 8.64E-03 | 6.95E-04 | 5.29E-05 | 7.45E-03 | 5.76E-04 | 4.36E-05 |
| | 0.05 | 0.01 | 1.47E-02 | 1.19E-03 | 7.48E-05 | 1.01E-02 | 7.19E-04 | 4.04E-05 |
| | 0.05 | 0.05 | 7.62E-03 | 5.85E-04 | 4.42E-05 | 6.38E-03 | 4.52E-04 | 3.38E-05 |
| | 0.05 | 0.5 | 6.03E-03 | 4.32E-04 | 3.31E-05 | 5.46E-03 | 3.83E-04 | 2.99E-05 |

Table S4: estimated Type 1 error rates in simulations for each of the BinomiRare and CMP tests, in each of the settings. The Type 1 error rates here were estimated using all simulations, including those in which the variance component was estimated as zero. In this case, CMP test reverts to the BinomiRare test. Orange colored cells correspond to settings in which the Type 1 error rates were higher than desired.

| Test | MAF | Odds | Mid-*p*-value Threshold: 1E-02 | Mid-*p*-value Threshold: 1E-03 | Mid-*p*-value Threshold: 1E-04 | p-value Threshold: 1E-02 | p-value Threshold: 1E-03 | p-value Threshold: 1E-04 |
|---|---|---|---|---|---|---|---|---|
| $\sigma_g^2 = 0.6$ | | | | | | | | |
| BinomiRare | 0.001 | 0.01 | 3.54E-03 | 3.63E-04 | 2.24E-05 | 2.05E-03 | 1.59E-04 | 9.05E-06 |
| | 0.001 | 0.05 | 5.71E-03 | 4.47E-04 | 3.22E-05 | 2.84E-03 | 2.16E-04 | 1.67E-05 |
| | 0.001 | 0.5 | 5.49E-03 | 2.99E-04 | 1.26E-05 | 2.61E-03 | 1.30E-04 | 5.30E-06 |
| | 0.01 | 0.01 | 6.63E-03 | 5.13E-04 | 3.80E-05 | 3.58E-03 | 2.70E-04 | 2.07E-05 |
| | 0.01 | 0.05 | 7.89E-03 | 6.62E-04 | 5.27E-05 | 5.08E-03 | 4.04E-04 | 2.86E-05 |
| | 0.01 | 0.5 | 8.25E-03 | 7.33E-04 | 6.61E-05 | 6.75E-03 | 5.71E-04 | 4.79E-05 |
| | 0.02 | 0.01 | 6.55E-03 | 4.82E-04 | 3.33E-05 | 3.73E-03 | 2.58E-04 | 1.71E-05 |
| | 0.02 | 0.05 | 7.62E-03 | 6.27E-04 | 5.04E-05 | 5.51E-03 | 4.27E-04 | 3.18E-05 |
| | 0.02 | 0.5 | 7.59E-03 | 6.38E-04 | 5.59E-05 | 6.56E-03 | 5.32E-04 | 4.53E-05 |
| | 0.05 | 0.01 | 5.30E-03 | 3.02E-04 | 1.32E-05 | 3.25E-03 | 1.70E-04 | 7.80E-06 |
| | 0.05 | 0.05 | 5.97E-03 | 4.28E-04 | 2.74E-05 | 4.84E-03 | 3.36E-04 | 2.10E-05 |
| | 0.05 | 0.5 | 5.71E-03 | 4.19E-04 | 3.38E-05 | 5.17E-03 | 3.69E-04 | 2.99E-05 |
| CMP | 0.001 | 0.01 | 2.26E-02 | 4.56E-03 | 3.49E-04 | 1.99E-02 | 2.17E-03 | 1.46E-04 |
| | 0.001 | 0.05 | 5.10E-02 | 5.59E-03 | 3.51E-04 | 2.90E-02 | 2.65E-03 | 1.55E-04 |
| | 0.001 | 0.5 | 6.35E-02 | 2.39E-03 | 3.23E-05 | 3.17E-02 | 7.42E-04 | 7.94E-06 |
| | 0.01 | 0.01 | 1.25E-02 | 1.26E-03 | 1.05E-04 | 7.38E-03 | 6.86E-04 | 5.49E-05 |
| | 0.01 | 0.05 | 1.78E-02 | 1.76E-03 | 1.54E-04 | 1.19E-02 | 1.10E-03 | 8.97E-05 |
| | 0.01 | 0.5 | 1.06E-02 | 8.66E-04 | 6.32E-05 | 8.63E-03 | 6.70E-04 | 4.70E-05 |
| | 0.02 | 0.01 | 8.98E-03 | 8.17E-04 | 7.00E-05 | 5.53E-03 | 4.70E-04 | 3.78E-05 |
| | 0.02 | 0.05 | 1.16E-02 | 1.15E-03 | 1.05E-04 | 8.68E-03 | 7.97E-04 | 6.69E-05 |
| | 0.02 | 0.5 | 8.60E-03 | 6.95E-04 | 5.35E-05 | 7.43E-03 | 5.77E-04 | 4.41E-05 |
| | 0.05 | 0.01 | 4.81E-03 | 3.87E-04 | 2.44E-05 | 3.29E-03 | 2.35E-04 | 1.32E-05 |
| | 0.05 | 0.05 | 7.10E-03 | 5.33E-04 | 3.86E-05 | 5.88E-03 | 4.14E-04 | 2.96E-05 |
| | 0.05 | 0.5 | 6.04E-03 | 4.34E-04 | 3.35E-05 | 5.48E-03 | 3.85E-04 | 3.04E-05 |

Table S5: estimated Type 1 error rates in simulations focusing in which the variance component was estimated as 0. Results are provided only for the BinomiRare and CMP test, because CMP is not used in these settings.

| | | | Mid-*p*-value | | | p-value | | |
|---|---|---|---|---|---|---|---|---|
| | | | Threshold: | | | Threshold: | | |
| Test | MAF | Odds | 1E-02 | 1E-03 | 1E-04 | 1E-02 | 1E-03 | 1E-04 |
| $\sigma_g^2 = 0.6$ | | | | | | | | |
| | 0.001 | 0.01 | 3.43E-03 | 4.13E-04 | 2.41E-05 | 1.99E-03 | 1.76E-04 | 8.88E-06 |
| | 0.001 | 0.05 | 5.88E-03 | 4.62E-04 | 3.18E-05 | 2.91E-03 | 2.19E-04 | 1.54E-05 |
| | 0.001 | 0.5 | 6.24E-03 | 3.87E-04 | 1.24E-05 | 3.04E-03 | 1.62E-04 | 3.55E-06 |
| | 0.01 | 0.01 | 6.76E-03 | 5.37E-04 | 4.01E-05 | 3.61E-03 | 2.82E-04 | 2.20E-05 |
| | 0.01 | 0.05 | 8.02E-03 | 6.69E-04 | 5.62E-05 | 5.14E-03 | 4.15E-04 | 2.94E-05 |
| | 0.01 | 0.5 | 8.91E-03 | 7.78E-04 | 7.54E-05 | 7.32E-03 | 6.05E-04 | 5.61E-05 |
| BinomiRare | 0.02 | 0.01 | 6.62E-03 | 4.86E-04 | 3.31E-05 | 3.76E-03 | 2.60E-04 | 1.74E-05 |
| | 0.02 | 0.05 | 7.82E-03 | 6.52E-04 | 4.58E-05 | 5.63E-03 | 4.43E-04 | 2.98E-05 |
| | 0.02 | 0.5 | 8.10E-03 | 6.96E-04 | 6.28E-05 | 7.00E-03 | 5.85E-04 | 5.23E-05 |
| | 0.05 | 0.01 | 5.34E-03 | 3.04E-04 | 1.24E-05 | 3.23E-03 | 1.70E-04 | 7.76E-06 |
| | 0.05 | 0.05 | 6.06E-03 | 4.31E-04 | 2.76E-05 | 4.89E-03 | 3.38E-04 | 2.14E-05 |
| | 0.05 | 0.5 | 6.24E-03 | 4.66E-04 | 4.02E-05 | 5.66E-03 | 4.16E-04 | 3.84E-05 |

### 3. Power estimates from simulations in which the variance component was estimated as non-zero.

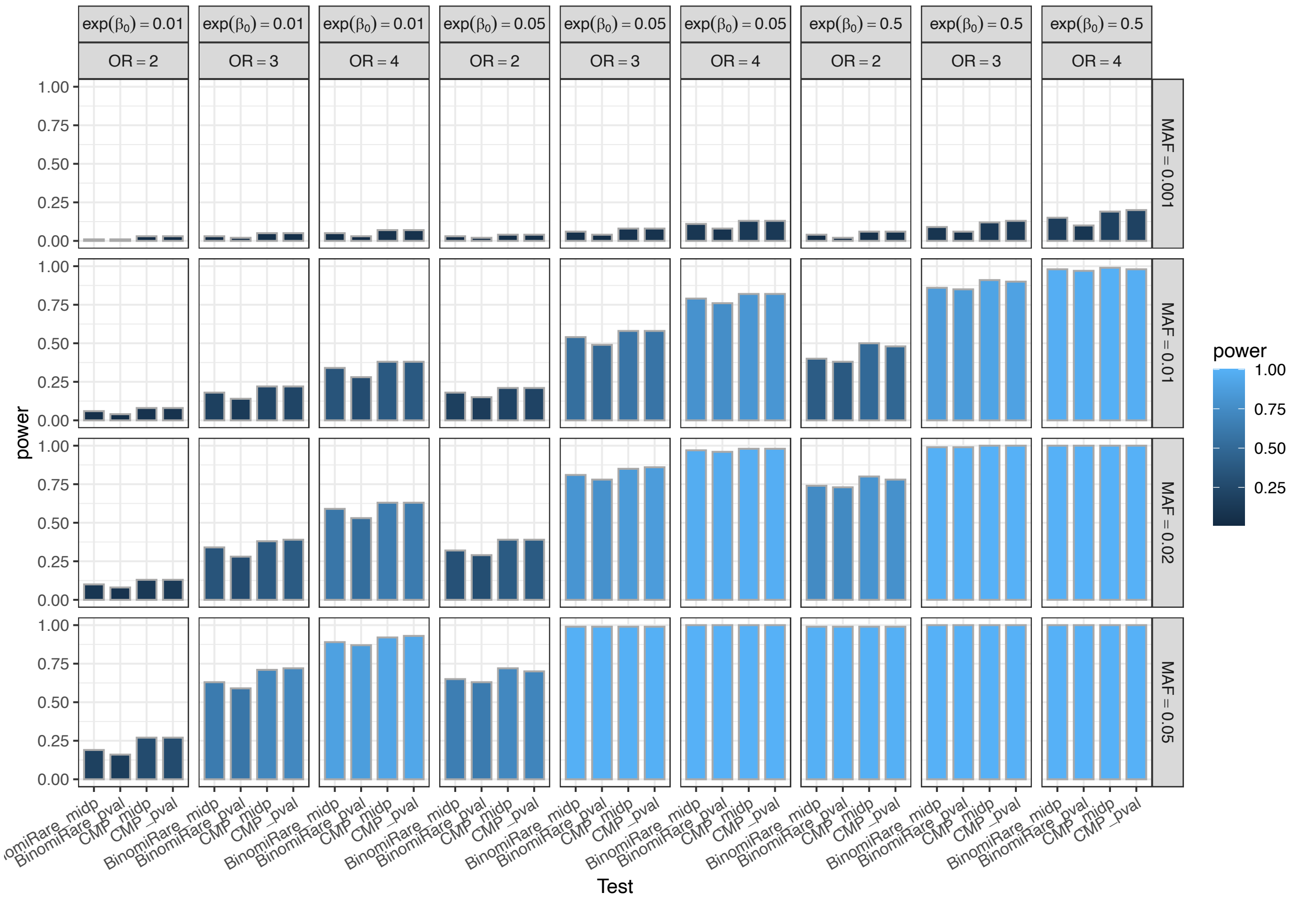

**Figure S1: Power estimates when testing using p-value threshold =0.01 ($\sigma_g^2 = 0.6$).**

Power estimates are based on simulations in which the variance component was estimated as non-zero. Patterns are similar when computing power on all simulations (including those in which variance components were estimated as zero). When CMP did had estimated Type 1 error rate larger than desired (the p-value threshold used for testing), we computed an “honest alpha” as the p-value threshold resulting in the desired Type 1 error, and used this threshold to compute power.

**Figure S2: Power estimates when testing using p-value threshold =1E-03 ($\sigma_g^2 = 0.6$).**

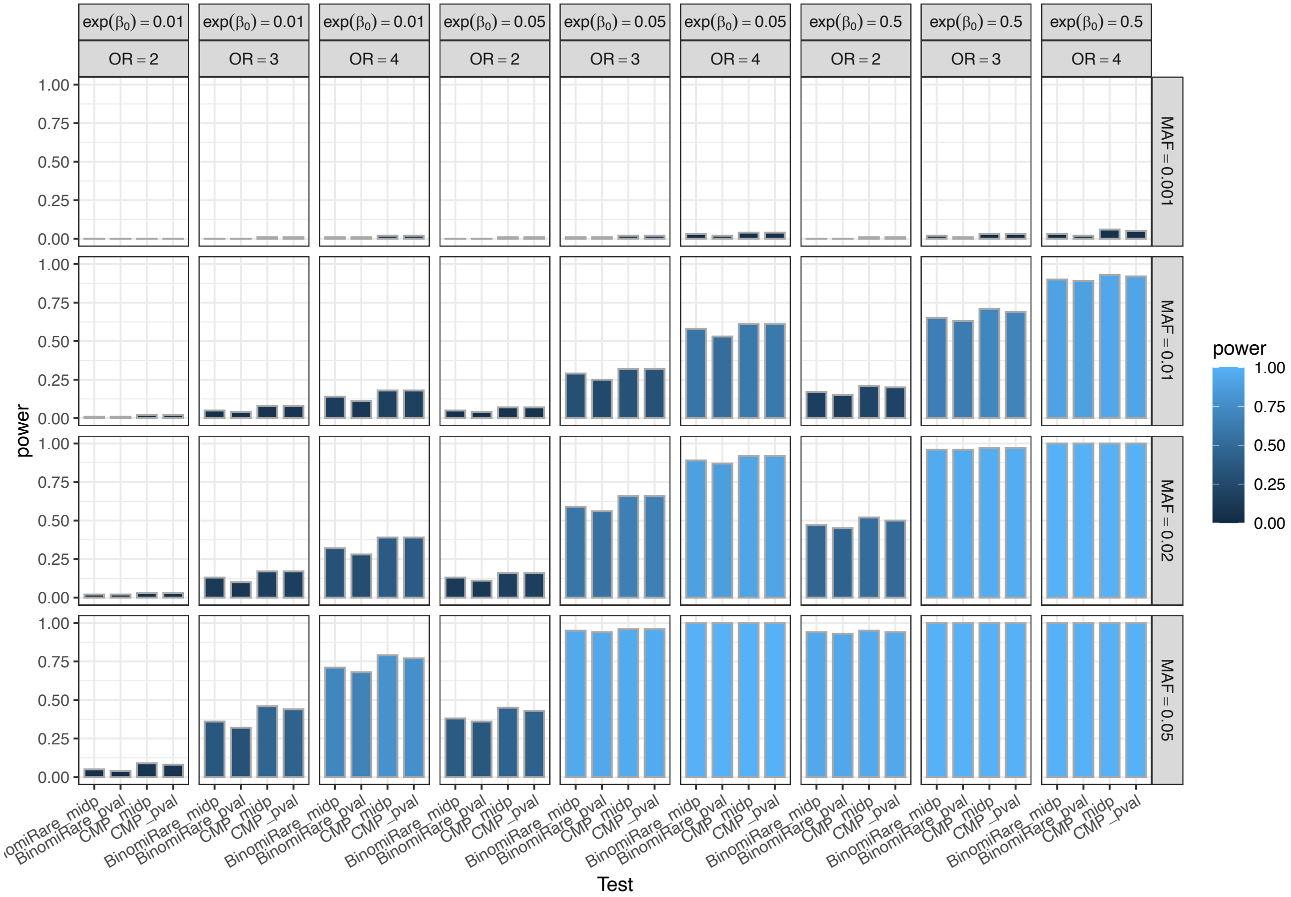

Power estimates are based on simulations in which the variance component was estimated as non-zero. Patterns are similar when computing power on all simulations (including those in which variance components were estimated as zero). When CMP did had estimated Type 1 error rate larger than desired (the p-value threshold used for testing), we computed an "honest alpha" as the p-value threshold resulting in the desired Type 1 error, and used this threshold to compute power.

**Figure S3: Power estimates when testing using p-value threshold =1E-04 ($\sigma_g^2 = 0.6$).**

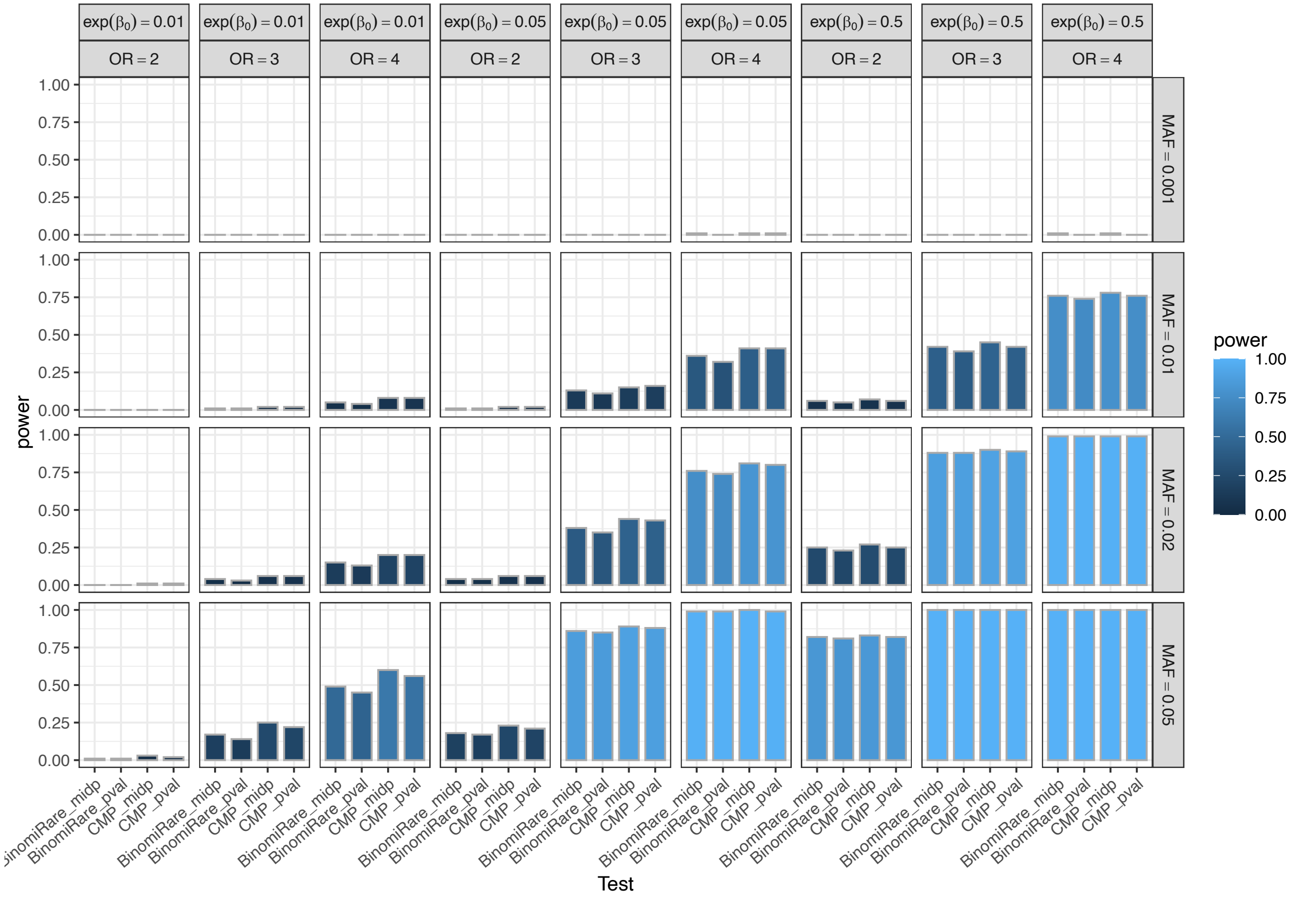

Power estimates are based on simulations in which the variance component was estimated as non-zero. Patterns are similar when computing power on all simulations (including those in which variance components were estimated as zero). When CMP did had estimated Type 1 error rate larger than desired (the p-value threshold used for testing), we computed an "honest alpha" as the p-value threshold resulting in the desired Type 1 error, and used this threshold to compute power.

### 4. Acknowledgements for participation studies.

**The Amish study**

We gratefully acknowledge our Amish liaisons, research volunteers, field workers and Amish Research Clinic staff and the extraordinary cooperation and support of the Amish community without which these studies would not have been possible. The Amish studies are supported by grants and contracts from the NIH, including U01 HL072515, U01 HL84756, U01 HL137181 and P30 DK72488. The TOPMed component of the Amish Research Program was supported by NIH grants R01 HL121007, U01 HL072515, and R01 AG18728. All study protocols were approved by the institutional review board at the University of Maryland Baltimore. Informed consent was obtained from each study participant. The Amish study participated in the analysis of short sleep. Some of the sleep durations were based on the Amish HAPI study (1).

**Atherosclerosis Risk in Communities (ARIC)**

The ARIC study is a population-based prospective cohort study of cardiovascular disease sponsored by the National Heart, Lung, and Blood Institute (NHLBI). ARIC included 15,792 individuals, predominantly European American and African American, aged 45-64 years at baseline (1987-89) and chosen by probability sampling from four US communities. Cohort members completed three additional triennial follow-up examinations, a fifth exam in 2011-2013, a sixth exam in 2016-2017, and a seventh exam in 2018-2019. The ARIC study has been described in detail previously (2). The ARIC study participated in the analysis of short sleep, and in the analysis of VTE.

The Atherosclerosis Risk in Communities study has been funded in whole or in part with Federal funds from the National Heart, Lung, and Blood Institute, National Institutes of Health, Department of Health and Human Services (contract numbers HHSN268201700001I, HHSN268201700002I, HHSN268201700003I, HHSN268201700004I and HHSN268201700005I). The authors thank the staff and participants of the ARIC study for their important contributions.

**Cardiovascular Health Study (CHS)**

The Cardiovascular Health Study (CHS) is a population-based cohort study initiated by the National Heart, Lung and Blood Institute (NHLBI) in 1987 to determine the risk factors for development and progression of cardiovascular disease (CVD) in older adults, with an emphasis on subclinical measures (3). The study recruited 5,888 adults aged 65 or older at entry in four U.S. communities and conducted extensive annual clinical exams between 1989-1999 along with semi-annual phone calls, events adjudication, and subsequent data analyses and publications. Additional data are collected by studies ancillary to CHS. In June 1990, four Field Centers (Sacramento, CA; Hagerstown, MD; Winston-Salem, NC; Pittsburgh, PA) completed the recruitment of 5201 participants. Between November 1992 and June 1993, an additional 687 African Americans were recruited using similar methods. Blood samples were drawn from all participants at their baseline examination and during follow-up clinic visits and DNA was subsequently extracted from available samples. CHS analyses were limited to participants with available DNA who consented to genetic studies. The baseline examinations consisted of a home interview and a clinic examination that assessed not only traditional risk factors but also measures of subclinical disease, including carotid ultrasound, echocardiography, electrocardiography, and pulmonary function. Between enrollment and 1998-99, participants were seen in the clinic annually, and contacted by phone at 6-month intervals to collect information about hospitalizations and potential cardiovascular events. Major exam components were repeated during annual follow-up examinations through 1999. Cranial MRI scans, retinal photography, and tests of endothelial function were added as new components. Standard protocols for the identification and adjudication of events were implemented during follow-up. The adjudicated events are CHD, angina, heart failure (HF), stroke, transient ischemic attack (TIA), claudication and mortality. Adjudication of cause of death continues using a streamlined protocol; adjudication of other events ended in June 2015. Deep venous thrombosis and pulmonary embolism events from baseline through 2001 were adjudicated in an ancillary study: the Longitudinal Investigation of Thromboembolism Etiology (LITE). Since 1999, participants have been contacted every 6 months by phone, primarily to ascertain health status and for events follow-up. The study was initially approved by institutional review boards at the Field Centers (Wake Forest, University of California – Davis, Johns Hopkins University, University of Pittsburgh), the Core Laboratory (University of Vermont) and at the Coordinating Center (University of Washington). The University of Washington now handles CHS Data Repository approvals. All CHS participants provided informed consent, and the study was approved by the Institutional Review Board [or ethics review committee] of University Washington. CHS participated in the short sleep analysis, and VTE analysis.

This research was supported by contracts HHSN268201200036C, HHSN268200800007C, HHSN268201800001C, N01HC55222, N01HC85079, N01HC85080, N01HC85081, N01HC85082, N01HC85083, N01HC85086, 75N920210000D, and grants U01HL080295 and U01HL130114 from the National Heart, Lung, and Blood Institute (NHLBI), with additional contribution from the National Institute of Neurological Disorders and Stroke (NINDS). Additional support was provided by R01AG023629 from the National Institute on Aging (NIA). A full list of principal CHS investigators and institutions can be found at CHS-NHLBI.org. The content is solely the responsibility of the authors and does not necessarily represent the official views of the National Institutes of Health.

**Cleveland Family Study**

The Cleveland Family Study (CFS) was designed to examine the genetic basis of sleep apnea in 2,534 African-American and European-American individuals from 356 families. Index probands with confirmed sleep apnea were recruited from sleep centers in northern Ohio, supplemented with additional family members and neighborhood control families (4).Four visits occurred between 1990 and 2006; in the first 3, data were collected in participants' homes while the last occurred in a clinical research center (2000 - 2006). Measurements included sleep apnea monitoring, blood pressure, anthropometry, spirometry and other related phenotypes. Blood samples (overnight fasting, before bed and following an oral glucose tolerance test), nasal and oral ultrasound, and ECG were also obtained during the 4th exam. Institutional Review Board approval and signed informed consent was obtained for all participants. Cleveland Family Study was approved by the Institutional Review Board (IRB) of Case Western Reserve University and

Mass General Brigham (formerly Partners HealthCare). Written informed consent was obtained from all participants. CFS participated in the analysis of short sleep.

The Cleveland Family Study has been supported in part by National Institutes of Health grants (R01-HL046380, KL2-RR024990, R35-HL135818, and R01-HL113338).

**Framingham Heart Study**

The Framingham Heart Study (FHS) acknowledges the support of contracts NO1-HC-25195 and HHSN268201500001I from the National Heart, Lung and Blood Institute and grant supplement R01 HL092577-06S1 for this research. We also acknowledge the dedication of the FHS study participants without whom this research would not be possible. The Framingham Heart Study was approved by the Institutional Review Board of the Boston University Medical Center. All study participants provided written informed consent. FHS participated in the analysis of short sleep and the analysis of VTE.

**Heart and Vascular Health Study (HVH)**

The Heart and Vascular Health (HVH) VTE Study is a case-control study of risk factors for cardiovascular outcomes set at Group Health (GH), an integrated health care delivery system in western Washington State. Cases include venous thromboembolism (VTE), myocardial infarction (MI), stroke, and atrial fibrillation (AF), with a shared common control group frequency matched to MI cases on age (within decade) sex, treated hypertension, and calendar year of identification (5). Study approval was granted by the human subjects committee at GH, and written informed consent was provided by all study participants. Eligibility and risk factor information were collected by trained medical record abstractors from a review of the GH medical record using only data available prior to the event date of cases and a randomly selected date for the controls. All VTE, MI, stroke and AF events were verified by medical record review. For the TOPMed data set, only incident idiopathic cases of VT and early-onset (age <=60 years) cases of AF without underlying heart failure, myocardial infarction, or valvular heart disease were included. Within the HVH study, VT and AF cases were diagnosed in both inpatient and outpatient settings. A venous blood sample was collected from all consenting subjects, and DNA was extracted from white blood cells using standard procedures. Study approval was granted by the human subjects committee at Group Health, and written informed consent was provided by all study participants. HVH participated in the VTE analysis

The Heart and Vascular Health Study was supported by grants HL068986, HL085251, HL095080, and HL073410 from the National Heart, Lung, and Blood Institute.

**Jackson Heart Study**

JHS is a longitudinal community-based study designed to assess the causes of the high prevalence of cardiovascular disease among AAs in the Jackson, Mississippi metropolitan area (6). During the baseline examination period (2000-2004) 5,306 self-identified AAs were recruited from urban and rural areas of the three counties (Hinds, Madison and Rankin) that comprise the Jackson, Mississippi metropolitan area. Participants were between 35 and 84 years old with the exception of a nested family cohort, where those ≥21 years old were eligible

All participants included in analyses provided written informed consent for genetic studies. Approval was obtained from the institutional review board of the University of Mississippi Medical Center (UMMC). Data on participants' health behaviors, medical history, and medication use were collected at baseline and subjects underwent venipuncture, allowing for assessment of complete blood cell counts and other measures at UMMC. The JHS study was approved by Jackson State University, Tougaloo College, and the University of Mississippi Medical Center IRBs, and all participants provided written informed consent. JHS participated in analysis of short sleep.

The Jackson Heart Study (JHS) is supported and conducted in collaboration with Jackson State University (HHSN268201300049C and HHSN268201300050C), Tougaloo College (HHSN268201300048C), and the University of Mississippi Medical Center (HHSN268201300046C and HHSN268201300047C) contracts from the National Heart, Lung, and Blood Institute (NHLBI) and the National Institute for Minority Health and Health Disparities (NIMHD). The authors also wish to thank the staffs and participants of the JHS.

**Mayo Clinic Venous Thromboembolism Study (Mayo_VTE)**

All Mayo_VTE participants provided informed consent and the study was approved by the Institutional Review Board of Mayo Clinic, Rochester, MN. Mayo_VTE participated in the VTE analysis.

The Mayo Venous Thromboembolism Study is funded in part, by grants from the National Institutes of Health, National Heart, Lung and Blood Institute (HL66216 and HL83141), the National Human Genome Research Institute (HG04735, HG06379), and research support provided by Mayo Foundation.

**Multi-Ethnic Study of Atherosclerosis (MESA)**

The Multi-Ethnic Study of Atherosclerosis is a study of the characteristics of subclinical cardiovascular disease (disease detected non-invasively before it has produced clinical signs and symptoms) and the risk factors that predict progression to clinically overt cardiovascular disease or progression of the subclinical disease (7). MESA consisted of a diverse, population-based sample of an initial 6,814 asymptomatic men and women aged 45-84. 38 percent of the recruited participants were white, 28 percent African American, 22 percent Hispanic, and 12 percent Asian, predominantly of Chinese descent. Participants were recruited from six field centers across the United States: Wake Forest University, Columbia University, Johns Hopkins University, University of Minnesota, Northwestern University and University of California - Los Angeles. Participants are being followed for identification and characterization of cardiovascular disease events, including acute myocardial infarction and other forms of coronary heart disease (CHD), stroke, and congestive heart failure; for cardiovascular disease interventions; and for mortality. The first examination took place over two years, from July 2000 - July 2002. It was followed by four examination periods that were 17-20 months in length. All MESA participants provided written informed consent, and the study was approved by the Institutional Review Boards at The Lundquist Institute (formerly Los Angeles BioMedical Research Institute) at Harbor-UCLA Medical Center, University of Washington, Wake Forest School of Medicine, Northwestern University, University of Minnesota, Columbia University, and Johns Hopkins University. MESA participated in the analysis of short sleep.

MESA and the MESA SHARe project are conducted and supported by the National Heart, Lung, and Blood Institute (NHLBI) in collaboration with MESA investigators. Support for MESA is provided by contracts HHSN268201500003I, N01-HC-95159, N01-HC-95160, N01-HC-95161, N01-HC-95162, N01-HC-95163, N01-HC-95164, N01-HC-95165, N01-HC-95166, N01-HC-95167, N01-HC-95168, N01-HC-95169, UL1-TR-000040, UL1-TR-001079, UL1-TR-001420. MESA Family is conducted and supported by the National Heart, Lung, and Blood Institute (NHLBI) in collaboration with MESA investigators. Support is provided by grants and contracts R01HL071051, R01HL071205, R01HL071250, R01HL071251, R01HL071258, R01HL071259, and by the National Center for Research Resources, Grant UL1RR033176. The provision of genotyping data was supported in part by the National Center for Advancing Translational Sciences, CTSI grant UL1TR001881, and the National Institute of Diabetes and Digestive and Kidney Disease Diabetes Research Center (DRC) grant DK063491 to the Southern California Diabetes Endocrinology Research Center.

**Women's Health Initiative (WHI)**

The WHI is a prospective national health study focused on identifying optimal strategies for preventing chronic diseases that are the major causes of death and disability in postmenopausal women (8). The WHI initially recruited 161,808 women between 1993 and 1997 with the goal of including a socio-demographically diverse population with racial/ethnic minority groups proportionate to the total minority population of US women aged 50-79 years. The WHI consists of two major parts: a set of randomized Clinical Trials and an Observational Study. The WHI Clinical Trials (CT; N=68,132) includes three overlapping components, each a randomized controlled comparison: the Hormone Therapy Trials (HT), Dietary Modification Trial, and Calcium and Vitamin D Trial. A parallel prospective observational study (OS; N = 93,676) examined biomarkers and risk factors associated with various chronic diseases. While the HT trials ended in the mid-2000s, active follow-up of the WHI-CT and WHI-OS cohorts has continued for over 25 years, with the accumulation of large numbers of diverse clinical outcomes, risk factor measurements, medication use, and many other types of data. All WHI participants provided informed consent and the study was approved by the Institutional Review Board (IRB) of the Fred Hutchinson Cancer Research Center. WHI participated in the stroke, VTE, and short sleep analyses.

The WHI program is funded by the National Heart, Lung, and Blood Institute, National Institutes of Health, U.S. Department of Health and Human Services through contracts HHSN268201600018C, HHSN268201600001C, HHSN268201600002C, HHSN268201600003C, and HHSN268201600004C.

### 5. TOPMed acknowledgements

Whole genome sequencing (WGS) for the Trans-Omics in Precision Medicine (TOPMed) program was supported by the National Heart, Lung and Blood Institute (NHLBI). WGS for "NHLBI TOPMed: Transomics for Precision Medicine Whole Genome Sequencing Project: ARIC" (phs001211.v3.p3) was performed at Baylor College of Medicine Human Genome Sequencing Center (HHSN268201500015C and 3U54HG003273-12S2) and the Broad Institute of MIT and Harvard (3R01HL092577-06S1). WGS for "NHLBI TOPMed: Genetics of Cardiometabolic Health in the Amish" (phs000956) was performed at the Broad Institute of MIT and Harvard (3R01HL121007-01S1). WGS for "NHLBI TOPMed: Cleveland Family Study - WGS Collaboration" (phs000954) was performed at the University of Washington Northwest Genomics Center (3R01HL098433-05S1, HHSN268201600032I). WGS for "NHLBI TOPMed: Cardiovascular Health Study" (phs001368.v2.p2) was performed at Baylor Genome Sequencing Center (3U54HG003273-12S2, HHSN268201500015C, HHSN268201600033I). WGS for "NHLBI TOPMed: Framingham Heart Study" (phs000974) was performed at the Broad Institute of MIT and Harvard (3R01HL092577-06S1, 3U54HG003067-12S2). WGS for "NHLBI TOPMed: Heart and Vascular Health Study (HVH)" (phs000993.v4.p2) was performed at Baylor Genome Sequencing Center (3U54HG003273-12S2, HHSN268201500015C). WGS for "NHLBI TOPMed: Jackson Heart Study" (phs000964) was performed at the University of Washington Northwest Genomics Center (HHSN268201100037C). WGS for "NHLBI TOPMed: Multi Ethnic Study of Atherosclerosis" (phs001416) was performed at the Broad Institute of MIT and Harvard (3R01HL092577-06S1, 3U54HG003067-12S2). WGS for "NHLBI TOPMed: Whole Genome Sequencing of Venous Thromboembolism (WGS or VTE)" (phs001402.v2.p1) was performed at

Baylor Genome Sequencing Center (3U54HG003273-12S2, HHSN268201500015C). WGS for "NHLBI TOPMed: Women's Health Initiative (WHI)" (phs001237.v2.p1) was performed at the Broad Institute of MIT and Harvard (HHSN268201500014C).

Centralized read mapping and genotype calling, along with variant quality metrics and filtering were provided by the TOPMed Informatics Research Center (3R01HL-117626-02S1; contract HHSN268201800002I). Phenotype harmonization, data management, sample-identity QC, and general study coordination were provided by the TOPMed Data Coordinating Center (3R01HL-120393-02S1; contract HHSN268201800001I). We gratefully acknowledge the studies and participants who provided biological samples and data for TOPMed.

### 6. CCDG acknowledgements

The Genome Sequencing Program (GSP) was funded by the National Human Genome Research Institute (NHGRI), the National Heart, Lung, and Blood Institute (NHLBI), and the National Eye Institute (NEI). The GSP Coordinating Center (U24 HG008956) contributed to cross-program scientific initiatives and provided logistical and general study coordination. The Centers for Common Disease Genomics (CCDG) program was supported by NHGRI and NHLBI, and whole genome sequencing was performed at the Baylor College of Medicine Human Genome Sequencing Center (UM1 HG008898 and R01HL059367).

### 7. TOPMed consortium authors

Namiko Abe[1], Goncalo Abecasis[2], Francois Aguet[3], Christine Albert[4], Laura Almasy[5], Alvaro Alonso[6], Seth Ament[7], Peter Anderson[8], Pramod Anugu[9], Deborah Applebaum-Bowden[10],

Kristin Ardlie[3], Dan Arking[11], Donna K Arnett[12], Allison Ashley-Koch[13], Stella Aslibekyan[14], Tim Assimes[15], Paul Auer[16], Dimitrios Avramopoulos[11], Najib Ayas[17], John Barnard[18], Kathleen Barnes[19], R. Graham Barr[20], Emily Barron-Casella[11], Lucas Barwick[21], Terri Beaty[11], Gerald Beck[18], Diane Becker[11], Lewis Becker[11], Rebecca Beer[22], Amber Beitelshees[7], Emelia Benjamin[23], Takis Benos[24], Marcos Bezerra[25], Larry Bielak[2], Joshua Bis[8], Thomas Blackwell[2], John Blangero[26], Eric Boerwinkle[27], Donald W. Bowden[28], Russell Bowler[29], Jennifer Brody[8], Ulrich Broeckel[30], Jai Broome[8], Deborah Brown[27], Karen Bunting[1], Esteban Burchard[31], Carlos Bustamante[15], Erin Buth[8], Brian Cade[32], Jonathan Cardwell[33], Vincent Carey[32], Julie Carrier[34], Cara Carty[35], Richard Casaburi[36], Juan P Casas Romero[32], James Casella[11], Peter Castaldi[32], Mark Chaffin[3], Christy Chang[7], Yi-Cheng Chang[37], Daniel Chasman[32], Sameer Chavan[33], Bo-Juen Chen[1], Wei-Min Chen[38], Yii-Der Ida Chen[39], Michael Cho[32], Seung Hoan Choi[3], Lee-Ming Chuang[37], Mina Chung[18], Ren-Hua Chung[40], Clary Clish[3], Suzy Comhair[18], Matthew Conomos[8], Elaine Cornell[41], Adolfo Correa[9], Carolyn Crandall[36], James Crapo[29], L. Adrienne Cupples[42], Joanne Curran[26], Jeffrey Curtis[2], Brian Custer[43], Coleen Damcott[7], Dawood Darbar[44], Sayantan Das[2], Sean David[45], Colleen Davis[8], Michelle Daya[33], Mariza de Andrade[46], Lisa de las Fuentes[47], Michael DeBaun[48], Ranjan Deka[49], Dawn DeMeo[32], Scott Devine[7], Qing Duan[50], Ravi Duggirala[26], Jon Peter Durda[41], Susan Dutcher[47], Charles Eaton[51], Lynette Ekunwe[9], Adel El Boueiz[52], Patrick Ellinor[53], Leslie Emery[8], Serpil Erzurum[18], Charles Farber[38], Tasha Fingerlin[29], Matthew Flickinger[2], Myriam Fornage[27], Nora Franceschini[50], Chris Frazar[8], Mao Fu[7], Stephanie M. Fullerton[8], Lucinda Fulton[47], Stacey Gabriel[3], Weiniu Gan[22], Shanshan Gao[33], Yan Gao[9], Margery Gass[54], Bruce Gelb[55], Xiaoqi (Priscilla) Geng[2], Mark Geraci[56], Soren Germer[1], Robert Gerszten[57], Auyon Ghosh[32], Richard Gibbs[58], Chris Gignoux[15], Mark Gladwin[24], David Glahn[59], Stephanie Gogarten[8], Da-Wei Gong[7], Harald Goring[26], Sharon Graw[19], Kathryn J. Gray[60], Daniel Grine[33], C. Charles Gu[47], Yue Guan[7], Xiuqing Guo[39], Namrata Gupta[3], David Haas[56], Jeff Haessler[54], Michael Hall[9], Daniel Harris[7], Nicola L. Hawley[61], Jiang He[62], Ben Heavner[8], Susan Heckbert[8], Ryan Hernandez[31], David Herrington[28], Craig Hersh[32], Bertha Hidalgo[14], James Hixson[27], Brian Hobbs[32], John Hokanson[33], Elliott Hong[7], Karin Hoth[63], Chao (Agnes) Hsiung[40], Yi-Jen Hung[64], Haley Huston[65], Chii Min Hwu[66], Marguerite Ryan Irvin[14], Rebecca Jackson[67], Deepti Jain[8], Cashell Jaquish[22], Min A Jhun[2], Jill Johnsen[65], Andrew Johnson[22], Craig Johnson[8], Rich Johnston[6], Kimberly Jones[11], Hyun Min Kang[2], Robert Kaplan[68], Sharon Kardia[2], Sekar Kathiresan[3], Shannon Kelly[43], Eimear Kenny[55], Michael Kessler[7], Alyna Khan[8], Wonji Kim[52], Greg Kinney[33], Barbara Konkle[65], Charles Kooperberg[54], Holly Kramer[69], Christoph Lange[70], Ethan Lange[33], Leslie Lange[33], Cathy Laurie[8], Cecelia Laurie[8], Meryl LeBoff[32], Jiwon Lee[32], Seunggeun Shawn Lee[2], Wen-Jane Lee[66], Jonathon LeFaive[2], David Levine[8], Dan Levy[22], Joshua Lewis[7], Xiaohui Li[39], Yun Li[50], Henry Lin[39], Honghuang Lin[42], Keng Han Lin[2], Xihong Lin[70], Simin Liu[51], Yongmei Liu[13], Yu Liu[15], Ruth J.F. Loos[55], Steven Lubitz[53], Kathryn Lunetta[42], James Luo[22], Ulysses Magalan[67], Michael Mahaney[26], Barry Make[11], Ani Manichaikul[38], JoAnn Manson[32], Lauren Margolin[3], Lisa Martin[71], Susan Mathai[33], Rasika Mathias[11], Susanne May[8], Patrick McArdle[7], Merry-Lynn McDonald[14], Sean McFarland[52], Stephen McGarvey[51], Daniel McGoldrick[8], Caitlin McHugh[8], Becky McNeil[72], Hao Mei[9], Luisa Mestroni[19], Deborah A Meyers[73], Emmanuel Mignot[15], Julie Mikulla[22], Nancy Min[9], Mollie Minear[22], Ryan L Minster[24], Braxton D. Mitchell[7], Matt Moll[32], May E. Montasser[7], Courtney Montgomery[74], Arden Moscati[55], Solomon Musani[9], Stanford Mwasongwe[9], Josyf C Mychaleckyj[38], Girish Nadkarni[55], Rakhi Naik[11], Take Naseri[75], Pradeep Natarajan[3], Sergei Nekhai[76], Sarah C. Nelson[8], Bonnie Neltner[33], Deborah Nickerson[8], Kari

North[50], Jeff O'Connell[7], Tim O'Connor[7], Heather Ochs-Balcom[77], Allan Pack[78], David T. Paik[15], Nicholette Palmer[28], James Pankow[79], George Papanicolaou[22], Cora Parker[72], Afshin Parsa[7], Juan Manuel Peralta[26], Marco Perez[15], James Perry[7], Ulrike Peters[54], Patricia Peyser[2], Lawrence S Phillips[6], Toni Pollin[7], Wendy Post[11], Julia Powers Becker[33], Meher Preethi Boorgula[33], Michael Preuss[55], Bruce Psaty[8], Pankaj Qasba[22], Dandi Qiao[32], Zhaohui Qin[6], Nicholas Rafaels[33], Laura Raffield[50], Vasan S. Ramachandran [42], D.C. Rao[47], Laura Rasmussen-Torvik[80], Aakrosh Ratan[38], Susan Redline[32], Robert Reed[7], Elizabeth Regan[29], Alex Reiner[81], Muagututiâ€˜a Sefuiva Reupena[82], Ken Rice[8], Stephen Rich[38], Dan Roden[48], Carolina Roselli[3], Jerome Rotter[39], Ingo Ruczinski[11], Pamela Russell[33], Sarah Ruuska[65], Kathleen Ryan[7], Ester Cerdeira Sabino[83], Danish Saleheen[20], Shabnam Salimi[7], Steven Salzberg[11], Kevin Sandow[39], Vijay G. Sankaran[52], Christopher Scheller[2], Ellen Schmidt[2], Karen Schwander[47], David Schwartz[33], Frank Sciurba[24], Christine Seidman[84], Jonathan Seidman[84], Vivien Sheehan[85], Stephanie L. Sherman[6], Amol Shetty[7], Aniket Shetty[33], Wayne Hui-Heng Sheu[66], M. Benjamin Shoemaker[48], Brian Silver[86], Edwin Silverman[32], Jennifer Smith[2], Josh Smith[8], Nicholas Smith[8], Tanja Smith[1], Sylvia Smoller[68], Beverly Snively[28], Michael Snyder[15], Tamar Sofer[32], Nona Sotoodehnia[8], Adrienne M. Stilp[8], Garrett Storm[33], Elizabeth Streeten[7], Jessica Lasky Su[32], Yun Ju Sung[47], Jody Sylvia[32], Adam Szpiro[8], Carole Sztalryd[7], Daniel Taliun[2], Hua Tang[15], Margaret Taub[11], Kent D. Taylor[39], Matthew Taylor[19], Simeon Taylor[7], Marilyn Telen[13], Timothy A. Thornton[8], Machiko Threlkeld[8], Lesley Tinker[54], David Tirschwell[8], Sarah Tishkoff[78], Hemant Tiwari[14], Catherine Tong[8], Russell Tracy[41], Michael Tsai[79], Dhananjay Vaidya[11], David Van Den Berg[87], Peter VandeHaar[2], Scott Vrieze[79], Tarik Walker[33], Robert Wallace[63], Avram Walts[33], Fei Fei Wang[8], Heming Wang[88], Karol Watson[36], Daniel E. Weeks[24], Bruce Weir[8], Scott Weiss[32], Lu-Chen Weng[53], Jennifer Wessel[56], Cristen Willer[2], Kayleen Williams[8], L. Keoki Williams[89], Carla Wilson[32], James Wilson[57], Quenna Wong[8], Joseph Wu[15], Huichun Xu[7], Lisa Yanek[11], Ivana Yang[33], Rongze Yang[7], Norann Zaghloul[7], Maryam Zekavat[3], Yingze Zhang[24], Snow Xueyan Zhao[29], Wei Zhao[2], Degui Zhi[27], Xiang Zhou[2], Xiaofeng Zhu[90], Michael Zody[1], Sebastian Zoellner[2]

1 - New York Genome Center; 2 - University of Michigan; 3 - Broad Institute; 4 - Cedars Sinai; 5 - Children's Hospital of Philadelphia, University of Pennsylvania; 6 - Emory University; 7 - University of Maryland; 8 - University of Washington; 9 - University of Mississippi; 10 - National Institutes of Health; 11 - Johns Hopkins University; 12 - University of Kentucky; 13 - Duke University; 14 - University of Alabama; 15 - Stanford University; 16 - University of Wisconsin Milwaukee; 17 - Providence Health Care; 18 - Cleveland Clinic; 19 - University of Colorado Anschutz Medical Campus; 20 - Columbia University; 21 - The Emmes Corporation; 22 - National Heart, Lung, and Blood Institute, National Institutes of Health; 23 - Boston University, Massachusetts General Hospital; 24 - University of Pittsburgh; 25 - FundaÃ§Ã£o de Hematologia e Hemoterapia de Pernambuco - Hemope; 26 - University of Texas Rio Grande Valley School of Medicine; 27 - University of Texas Health at Houston; 28 - Wake Forest Baptist Health; 29 - National Jewish Health; 30 - Medical College of Wisconsin; 31 - University of California, San Francisco; 32 - Brigham & Women's Hospital; 33 - University of Colorado at Denver; 34 - University of Montreal; 35 - Washington State University; 36 - University of California, Los Angeles; 37 - National Taiwan University; 38 - University of Virginia; 39 - Lundquist Institute; 40 - National Health Research Institute Taiwan; 41 - University of Vermont; 42 - Boston University; 43 - Vitalant Research Institute; 44 - University of Illinois at Chicago; 45 -

University of Chicago; 46 - Mayo Clinic; 47 - Washington University in St Louis; 48 - Vanderbilt University; 49 - University of Cincinnati; 50 - University of North Carolina; 51 - Brown University; 52 - Harvard University; 53 - Massachusetts General Hospital; 54 - Fred Hutchinson Cancer Research Center; 55 - Icahn School of Medicine at Mount Sinai; 56 - Indiana University; 57 - Beth Israel Deaconess Medical Center; 58 - Baylor College of Medicine Human Genome Sequencing Center; 59 - Boston Children's Hospital, Harvard Medical School; 60 - Mass General Brigham; 61 - Yale University; 62 - Tulane University; 63 - University of Iowa; 64 - Tri-Service General Hospital National Defense Medical Center; 65 - Blood Works Northwest; 66 - Taichung Veterans General Hospital Taiwan; 67 - Oklahoma State University Medical Center; 68 - Albert Einstein College of Medicine; 69 - Loyola University; 70 - Harvard School of Public Health; 71 - George Washington University; 72 - RTI International; 73 - University of Arizona; 74 - Oklahoma Medical Research Foundation; 75 - Ministry of Health, Government of Samoa; 76 - Howard University; 77 - University at Buffalo; 78 - University of Pennsylvania; 79 - University of Minnesota; 80 - Northwestern University; 81 - Fred Hutchinson Cancer Research Center, University of Washington; 82 - Lutia I Puava Ae Mapu I Fagalele; 83 - Universidade de Sao Paulo; 84 - Harvard Medical School; 85 - Baylor College of Medicine; 86 - UMass Memorial Medical Center; 87 - University of Southern California; 88 - Brigham & Women's Hospital, Mass General Brigham; 89 - Henry Ford Health System; 90 - Case Western Reserve University
